# Exploring pathways to malaria control and elimination in the northern regions (Atacora, Alibori, Borgou, Donga) of Benin Republic: a metapopulation modelling approach

**DOI:** 10.1101/2025.07.09.25331180

**Authors:** Kounoumi Sara Medekon, Jules Degila, Sheetal Silal

## Abstract

**Background:** Malaria remains a major public health issue in sub-Saharan Africa, particularly in Benin, where it is the leading cause of morbidity and mortality. Despite ongoing control efforts, the disease burden remains high, necessitating a more comprehensive approach to explore differents pathways to malaria control in the northern regions (Atacora, Alibori, Borgou, Donga) of Benin Republic. This study employs a metapopulation mathematical model to assess the impact of key interventions, including insecticide-treated nets (ITNs), seasonal malaria chemoprevention (SMC), intermittent preventive treatment in pregnancy (IPTp), and indoor residual spraying (IRS) and effective anti-malaria treatment in the northern regions of Benin.

**Methods:** A metapopulation model stratified by age groups was developed to simulate malaria transmission dynamics in four highly endemic regions: Atacora, Alibori, Borgou, and Donga. The model incorporates seasonal variations, vector control measures, and treatment interventions. Calibration was performed using malaria incidence and prevalence data of Malaria Atlas Project from 2010 to 2022. Various intervention scenarios were simulated to determine their potential impact on malaria control.

**Results:** Among the intervention scenarios tested, the combination of all five strategies ITNs, SMC, IPTp, IRS, and treatment of malaria cases was predicted to be the most effective, reducing malaria incidence to a low level. In addition, scaling up ITN coverage in combination with effective anti-malaria treatment emerged as the second most effective strategy, reducing malaria cases, though it was less impactful than the comprehensive approach involving all five interventions. Other strategies, such as ITNs alone or combined with SMC also showed significant reductions in malaria burden particularly among children.

## Background

Malaria remains a major global health issue, with an estimated 263 million cases and an incidence of 60.4 cases per 1000 in 2023 [1]. The African Region continues to bear the highest burden, accounting for approximately 94% of all malaria cases worldwide in 2023 [1]. Furthermore, the disease remains the leading cause of child mortality in the continent. Nearly every minute, a child under 5 dies of malaria in Africa[2]. The burden is disproportionately felt in Sub-Saharan Africa, which accounted for approximately 94% of all reported malaria cases and deaths in 2023 [1]. In addition to its high mortality, malaria causes severe morbidity, particularly in children and pregnant women [1].

The disease is also a key socio-economic challenge, significantly hampering the growth and development of endemic countries, where billions of dollars are spent annually on combating the disease [3, 4]. As a matter of fact, Malaria imposes an estimated annual economic burden of approximately 12 billion dollars, representing a 1.3% reduction in GDP growth across malaria-endemic nations [1]. Families in malaria-endemic regions spend a significant portion of their income on treating and preventing malaria, which hinders overall economic development [5]. The costs on the government per case of severe malaria treatment, adjusted to 2023 values, ranged from USD 27 in Uganda to USD 165 in Kenya [6]. Moreover, the high incidence of malaria-related morbidity and mortality also contributes to the burden on the health system, with malaria accounting for a large proportion of hospital admissions and medical visits [1, 7].

Benin, a country in West Africa, is among the nations most affected by malaria [7]. According to the National Malaria Control Program (NMCP) of Benin, nearly every individual in the country is at risk of contracting malaria [7]. In 2023, malaria has been declared as the highest ranking cause of death in Benin [1]. Benin’s malaria statistics reveal alarming figures: the country accounts for 2% of malaria cases in African Region [1]. In 2023, 5.2 million cases were reported, and 9,673 deaths occurred as a result of the disease in the country [1].

In the northern regions of Benin, including Atacora, Alibori, Donga, and Borgou illustrated in Figure 1, Malaria is highly prevalent [7, 8].

**Fig. 1:**
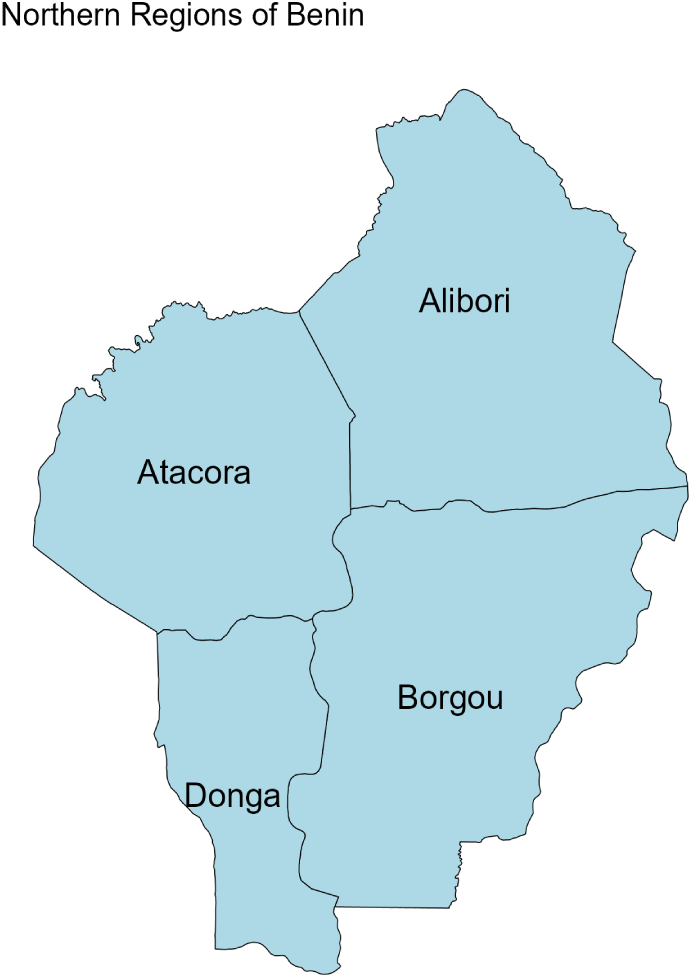
The four regions in the northern Benin

In response to this crisis, the government of Benin has committed to malaria elimination by 2030, as outlined in the Integrated Strategic Plan for Malaria Elimination (PSNIE) 2020-2024. Several key interventions have been implemented to reduce the burden of malaria in the country [1, 8]. The National Malaria Control Programme (NMCP) of Benin has carried out multiple distributions of Long-Lasting Insecticidal Nets (LLINs) to expand coverage and reduce mosquito bites and kill mosquitoes, thereby lowering malaria transmission. Since 2010, with support from funders, homes and buildings particularly in northern Benin have been sprayed with insecticides to kill mosquitoes, protecting millions of people. In addition, pregnant women in Benin receive prophylatic treatment at regular intervals to reduce their risk of malaria, ensuring the health of both mother and child. In the northern regions, where malaria is highly seasonal, young children under five are provided with preventive medication during peak transmission periods to protect them from infection. The country is also committed to early diagnosis and prompt treatment of malaria cases, especially among high-risk groups. This approach not only helps reduce the severity and duration of illness but also lowers parasite levels in the population, contributing to reduced transmission.

Several studies have contributed to a growing understanding of malaria dynamics and control strategies in Benin, each addressing different aspects of transmission, intervention effectiveness, and environmental determinants. Moiroux et al. [9] and Moiroux et al. [10] developed high-resolution predictive maps of vector abundance and biting risk in southern Benin, revealing strong seasonal and spatial heterogeneity. Similarly, Boussari et al. [11] employed latent class trajectory modeling to classify villages based on biting rate profiles linked to ecological factors, supporting spatially targeted vector control. Environmental influences were further explored by Cottrell et al. [12], who showed how local environmental conditions drive variations in vector density, and by Arab et al. [13], who identified negative associations between malaria incidence and climate factors in Benin. In terms of interventions, Sovi et al. [14] highlighted increasing insecticide resistance in Anopheles mosquitoes post-LLIN campaigns, emphasizing the urgency of resistance management. Boussari et al. [15] evaluated combinations of LLINs, IRS, and CTPS (Carbamate-Treated Plastic Sheeting), showing mixed results in vector density reduction but identifying key environmental correlates. On the health system side, Keating et al. [16] reported generally good diagnostic performance, though gaps in testing and treatment practices were noted. Lastly, Camponovo et al. [17] used agent-based modeling to demonstrate that scaling up effective treatment coverage could lead to substantial reductions in malaria prevalence and incidence, particularly among young children. Together, these studies highlight the importance of spatial, environmental, and operational factors in shaping malaria transmission and the response landscape in Benin.

Widely used by researchers, compartmental models serve as powerful tools for understanding malaria transmission dynamics and assessing the effectiveness of various control interventions [18–35]. A metapopulation model, which divides a region into smaller subpopulations (or patches), is particularly useful for studying malaria dynamics [36–38] in areas with distinct ecological and demographic characteristics, such as the northern regions of Benin.

These models can be used to test various ”what-if” scenarios, such as increased LLIN coverage or combined interventions, to predict their potential impact on malaria incidence and prevalence.

The purpose of this paper is to assess the impact of malaria control interventions on the incidence of malaria (estimate of clinical cases per 1000 among children under five and estimate of reported treated cases per 1000) in Benin, specifically in the northern Benin (Atacora, Alibori, Donga, and Borgou). In particular, the research focuses firstly on assessing the impact of current interventions on malaria incidence in these regions and simulating different intervention scenarios, such as increasing ITN coverage or combining multiple interventions, like treatment and IRS, to explore likely pathways for achieving malaria elimination.

The results of this study will provide simulation-based insights for policymakers and health authorities in Benin, providing directions on interventions choices in these regions of the northern part of the country.

## 1 Methods

### 1.1 Assessing elimination of malaria

Elimination is defined as achieving and maintaining zero locally acquired malaria cases in a specific region for three consecutive years, in line with the WHO’s definition [1, 39]. In the model, an elimination threshold of 0.1/1000 is assumed. Once sustained zero local transmission is achieved over three years, the region is considered to have eliminated malaria.

### 1.2 Transmission model

A metapopulation model is developed to describe the dynamics of malaria transmission across different age groups, including children under 2 years, children aged 2 to 5 years, children aged 5 to 10 years, and adults. It is structured to incorporate different subpopulations, key epidemiological processes, and interventions in a metapopulation framework for Benin. The subpopulations are the four regions in the northern part of Benin where malaria cases are high in the country: Atacora, Alibori, Borgou and Donga. The model incorporates the effects of seasonal malaria chemoprevention (SMC) and intermittent preventive treatment in pregnancy (IPTp), indoor residual spraying (IRS), Insecticide-treated bed nets (ITNs) and effective treatment with an antimalarial medicine as interventions. Each age group is further divided into compartments representing different stages of the disease. In the model, we have for each age group Susceptible; Exposed; Asymptomatic; Clinical non treated; Clinical treated; Severe; Hospitalized; Treated; Recovered. For children under the age of five, there is a ’Protected by SMC’ compartment, which they enter each year from July to October. Similarly, pregnant women are protected by IPTp during the six months of pregnancy. A complete description of the model structure, parameter values, compartment names and definitions, as well as the underlying assumptions, is provided in the supplementary file.

### 1.3 Model flow

The children model is stratified into three age groups: under 2 years (see Figure 2), 2 to 5 years, and 5 to 10 years. Individuals enter the system through birth into the under-2 age group. As they age, they transition to the 2–5 years group and subsequently to the 5–10 years group, according to age-specific aging rates. Each age group follows a compartmental structure capturing the dynamics of malaria infection, including susceptible, infected, recovered, and treated states, as well as intervention effects such as SMC depending on age. The progression through age groups ensures that demographic changes are accurately reflected in disease dynamics and intervention impact within the pediatric population. The adult population (see Figure 3) is divided into three distinct subgroups to reflect varying levels of immunity and exposure: newly aged-in adults (Adult), first level immune adults (Adult 1), second level immune adults (Adult 2) and third level immune adults (Adult 3). Individuals enter the adult model from the children’s model at age 10, entering initially into the Adult compartment. Over time, individuals transition sequentially from Adult 1 to Adult 2 and then to Adult 3 based on the number of malaria episodes experienced, reflecting the gradual development of partial and eventually full immunity through repeated exposure to the parasite. Each adult subgroup includes compartments for susceptible, exposed, infected, recovered, and treated individuals, and incorporates intervention strategies such as insecticide-treated nets (ITNs), indoor residual spraying (IRS), IPTp and treatment of clinical cases. The structure captures both epidemiological transitions and the natural history of immunity development, providing a comprehensive representation of malaria dynamics among adults.

**Fig. 2:**
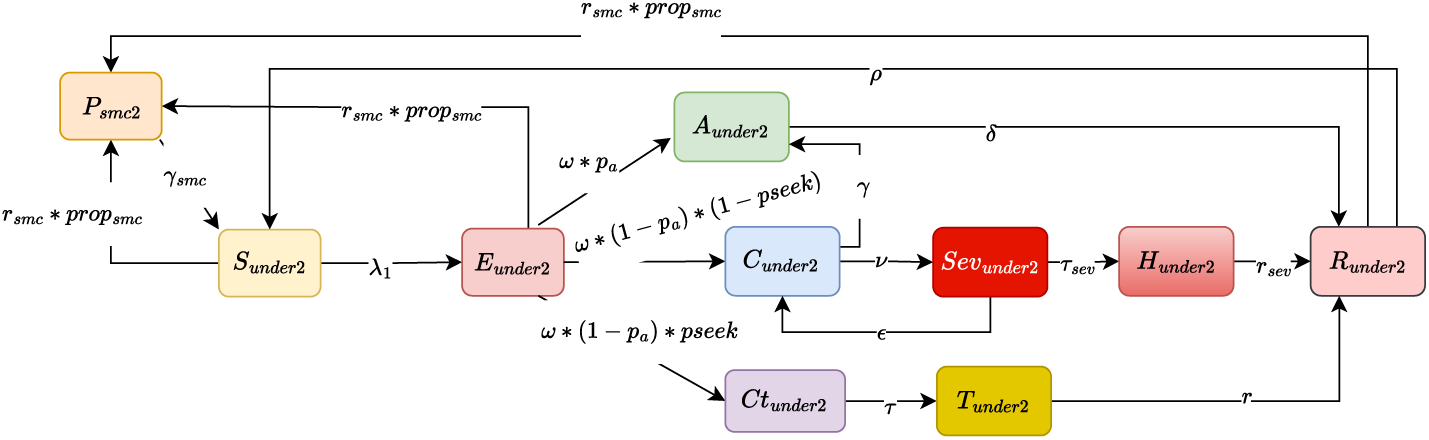
Model of transmission for children under two

**Fig. 3:**
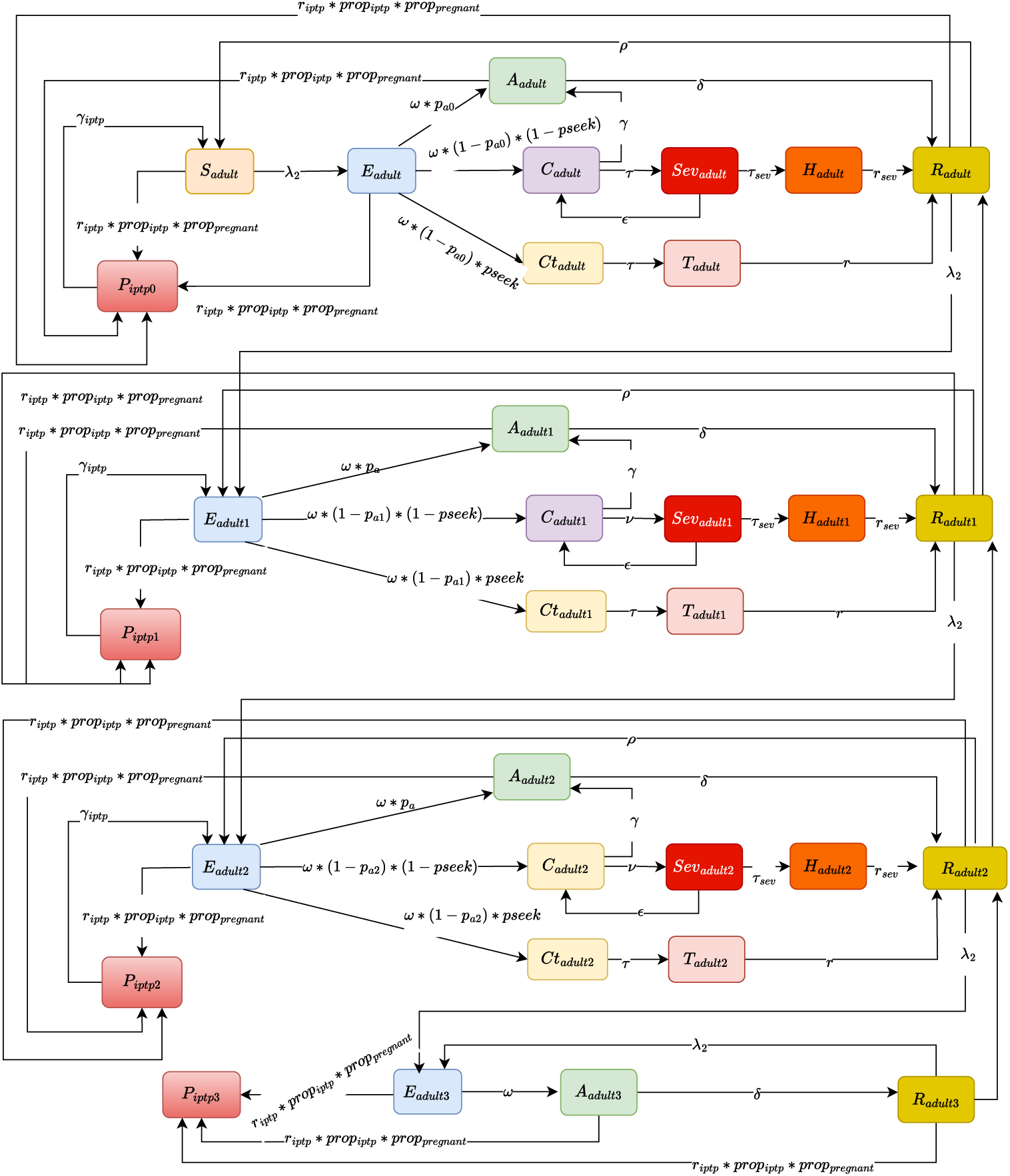
Model of transmission for adults and pregnant women with the three levels of immunity

A full diagram and description of model structure, parameters values and additional assumptions are available in the supplementary file. The model is implemented in R, and numerical integration are performed using the deSolve package.

### 1.4 Data

To inform and calibrate the model, spatially estimates of malaria trends data were obtained from the Malaria Atlas Project (MAP) for the northern region of Benin, covering the period from 2010 to 2022. Two key epidemiological indicators were extracted: malaria incidence (annual clinical cases per 1,000 population) and parasite prevalence (proportion of infected children aged 2–10 years).

Figures 4 and 5 present the spatial distribution and temporal trends of malaria incidence and prevalence across the four northern departments of Benin. Both indicators exhibit a substantial decline over time, indicating the effect of intensified control efforts. Nevertheless, spatial disparities persist, with some areas maintaining higher transmission intensities, particularly in earlier years.

**Fig. 4:**
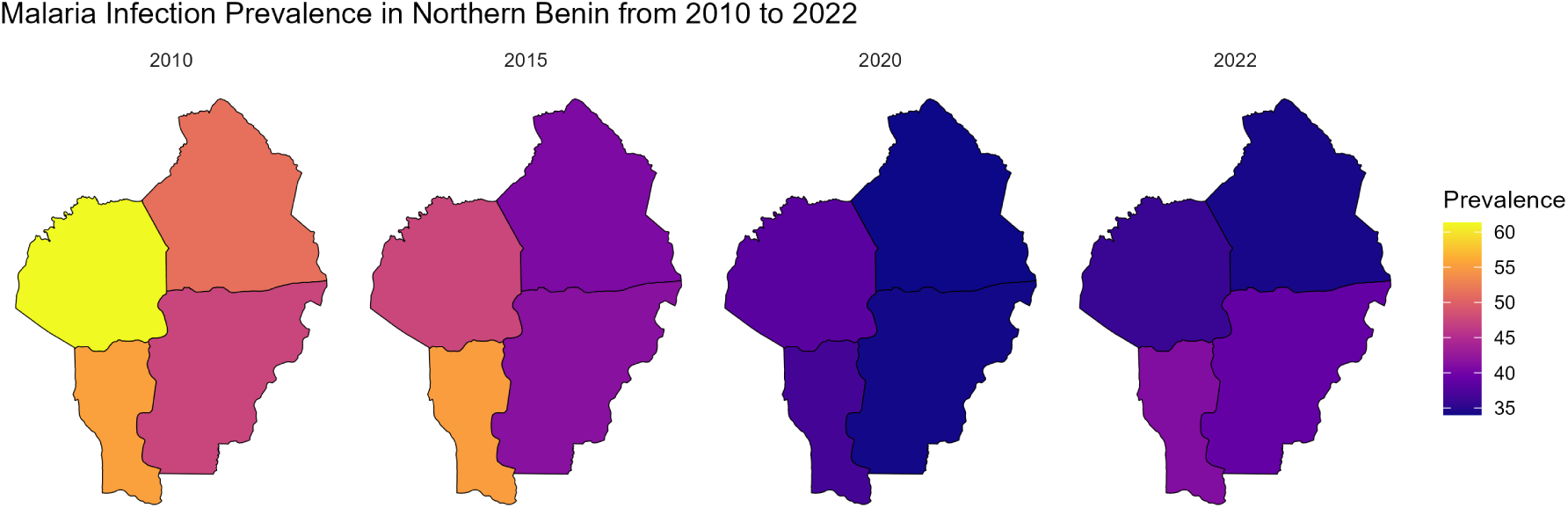
The proportion of children 2 to 10 years with *Plasmodium Falciparum* infection in Northern Benin from 2010 to 2022 from [40]

**Fig. 5:**
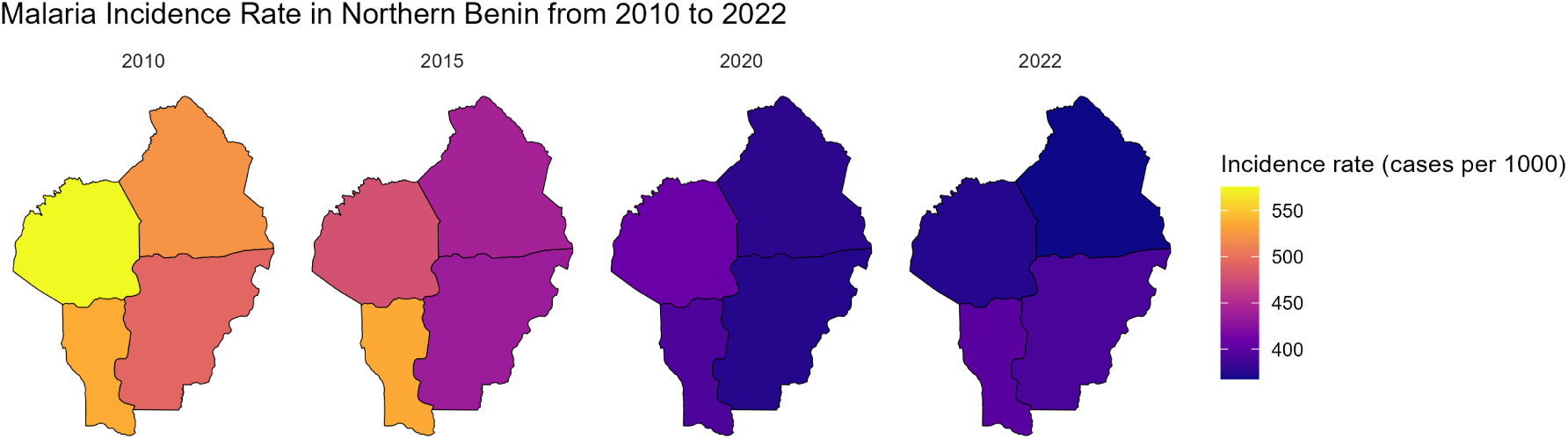
Malaria Incidence rate per 1000 in Northern Benin from 2010 to 2022 from [40]

These estimates from MAP provide a robust empirical foundation for the model fitting described in the following section, ensuring that the simulations are grounded in realistic and region-specific malaria dynamics.

The following Figures 6 and 8 illustrate the estimated coverage of key malaria interventions: Indoor Residual Spraying (IRS), Insecticide-Treated Nets (ITNs) and effective Treatment with Anti-malarial medicine from 2010 to 2022. These estimates are also obtained from the Malaria Atlas Project [40] and provide a regional overview of the scale-up and variation in intervention coverage over time in the northern regions of Benin. The data was crucial for informing the model and understanding the estimates trends in malaria control efforts across the study areas.

**Fig. 6:**
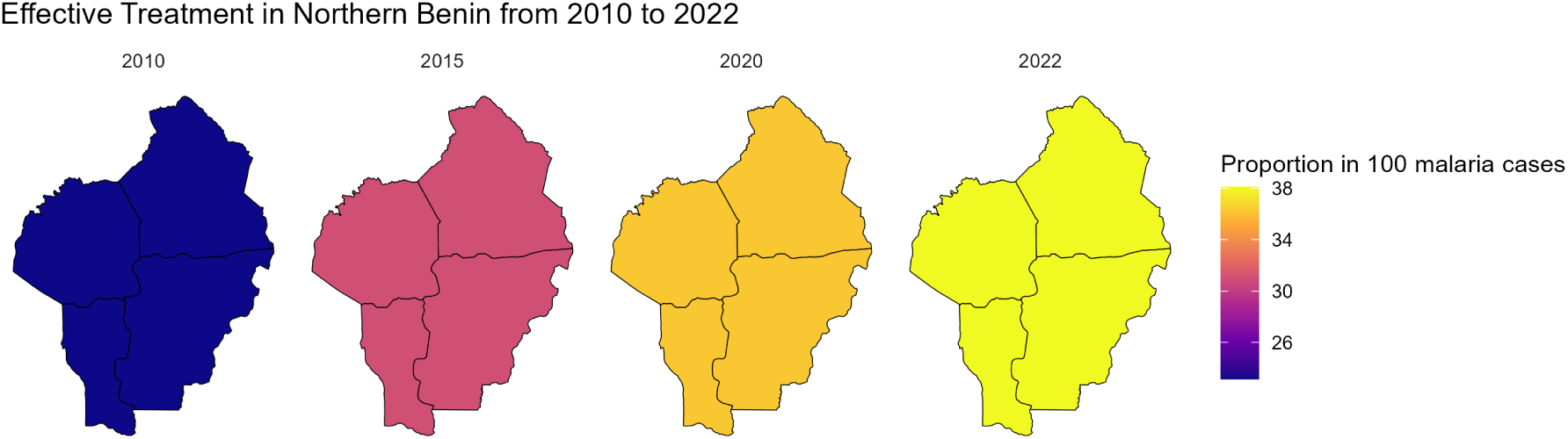
Proportion of malaria cases that receive effective Treatment with Anti-malarial medicine, estimates obtained from Malaria Atlas Project [40]

**Fig. 7:**
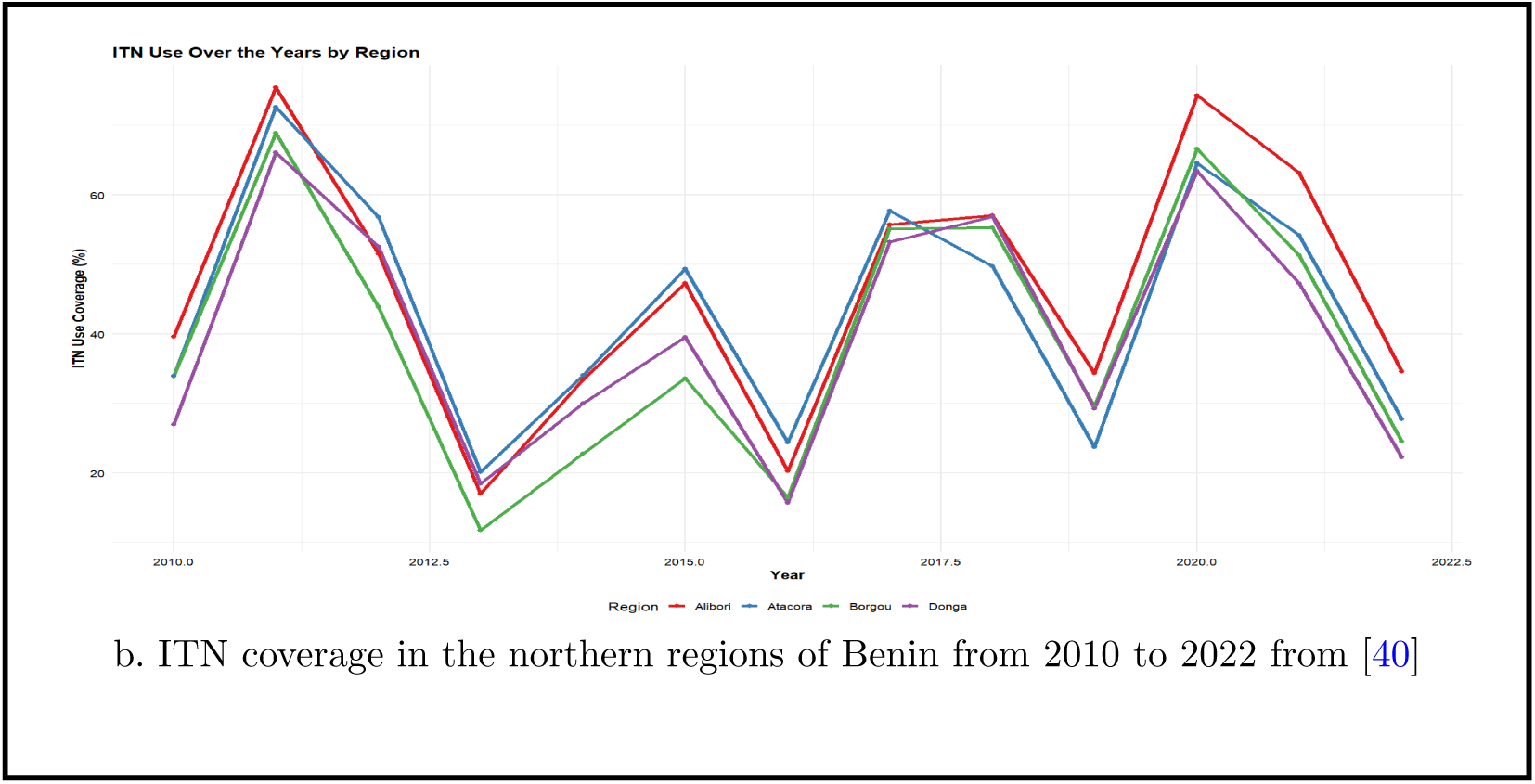
ITN coverage in the northern regions of Benin from 2010 to 2022 from [40]

**Fig. 8:**
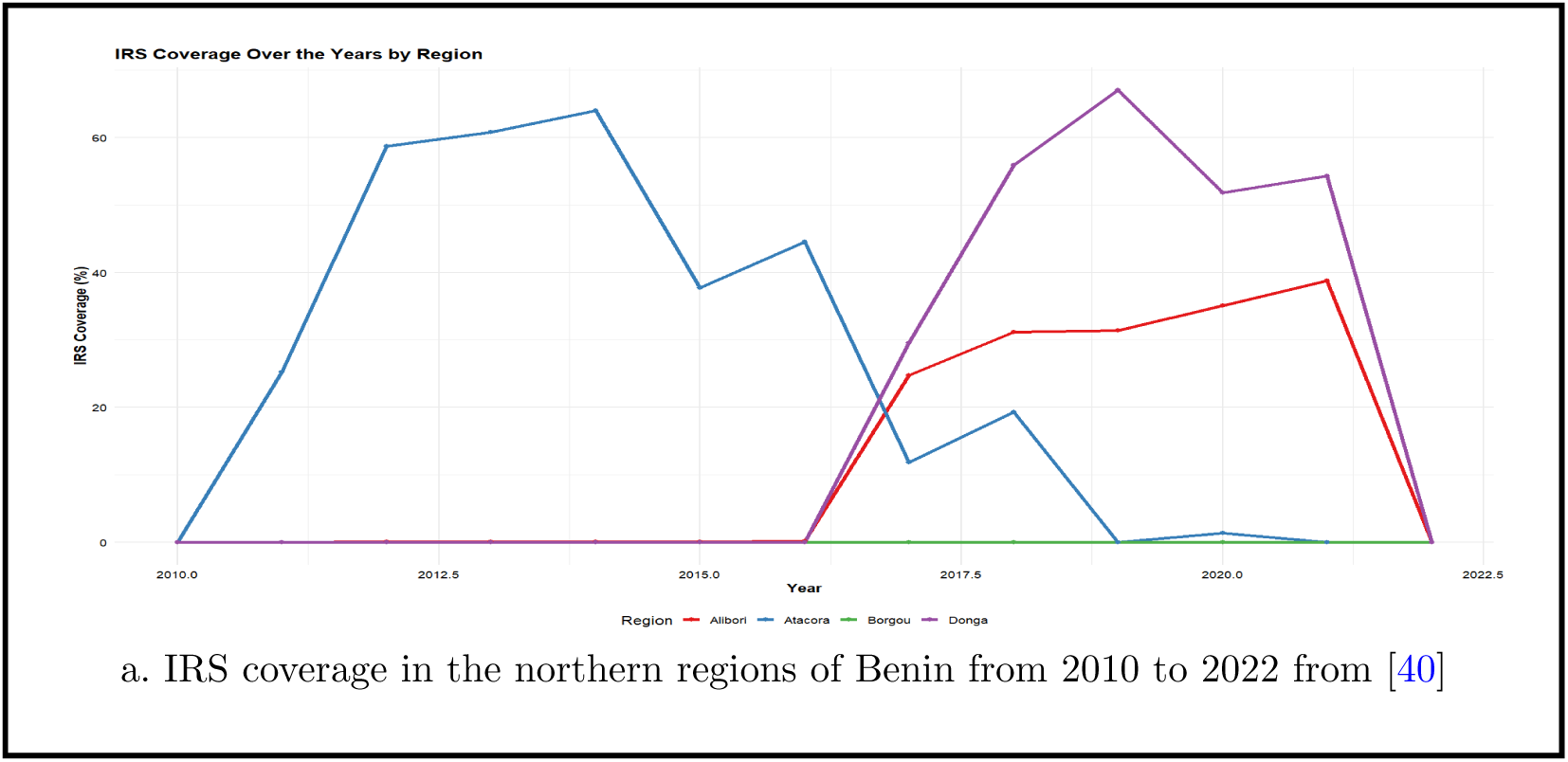
IRS coverage in the northern regions of Benin from 2010 to 2022 from [40]

### 1.5 Interventions

#### Interventions Modeled

The interventions modeled in this study aim to assess the impact of different malaria control strategies on the incidence of malaria in the northern regions of Benin. These interventions are simulated with varying levels of coverage, with a focus on achieving malaria elimination by 2030. The interventions include:

- Scenario 0: Baseline (business as usual):
- **Scenario 1: Scaling up ITN coverage and full treatment of malaria cases coverage**: This scenario simulates the combined effect of increasing ITN coverage and improving case management. Early diagnosis and complete treatment with effective antimalarial drugs are crucial for reducing malaria-related morbidity and mortality. ITN coverage are simulated to be increased to 70% in 2025 and follow an increase of 10% in 2026 and 2027 and treatment seeking rate is simulated to be increased to 50% in 2025 in the four regions and follow and increase of 10% in 2026 and 2027 to explore the impact of strengthening case management alongside vector control.
- **Scenario 2: Scaling up ITN**: In this scenario, we model the effect of increasing coverage of ITNs. The coverage of ITNs are simulated to be increased to 70% and follow an increase of 10% in 2026 and 2027.
- **Scenario 3: Scaling up ITN and SMC coverage**: This scenario models the combined effect of increasing coverage of long-lasting insecticidal nets (ITNs) and seasonal malaria chemoprevention (SMC) over three years from 2025 to 2027. ITNs provide a protective barrier against mosquito bites, while SMC, involving intermittent administration of antimalarial drugs to children during high transmission seasons, reduces malaria morbidity and mortality. The coverage of ITNs are simulated to be increased to 70% and follow an increase of 10% in 2026 and 2027. The coverage of SMC are simulated to be increased to 80% and follow an increase of 5% in 2026. This scenario evaluates the potential impact on malaria prevalence and incidence in the northern regions of Benin.
- **Scenario 5: Scaling up ITN, SMC, IPTp, IRS, and full treatment**: This scenario examines the combined effect of scaling up all five interventions: ITNs, SMC, IPTp, IRS, and full treatment of malaria cases. IRS coverage are simulated to be increased in areas where the coverage is below 50%, reaching 50% by 2027 and maintaining that level. It models the synergistic impact of improving vector control, preventive treatment during pregnancy, seasonal malaria chemoprevention for children, and ensuring case management. IRS coverage are simulated to gradually increase to 50%, and remains at these levels after. The coverage of ITNs are simulated to be increased to 70% and follow an increase of 10% in 2026 and 2027. The coverage of IPTp are simulated to be increased to 60% and follow an increase of 10% in 2026 and 2027 and treatment seeking rate is simulated to be increased to 50% in 2025 in the four regions and follow and increase of 10% in 2026 and 2027. Finally, The coverage of SMC are simulated to be increased to 80% and follow an increase of 5% in 2026.

Each of these intervention scenarios will be compared to determine the most effective strategy for reducing malaria incidence (estimate of clinical cases per 1000 among children under five and estimate of reported treated cases per 1000) in northern Benin.

**Table 1:** Coverage estimates for malaria interventions in 2021 [40, 41].

| Estimate Coverage in 2021 | Alibori | Atacora | Borgou | Donga |
| --- | --- | --- | --- | --- |
| IRS (%) | 38 | 1 | 0 | 54 |
| ITN (%) | 35 | 30 | 28.63 | 28 |
| SMC (%) | 70 | 70 | 0 | 0 |
| IPTp (%) | 50 | 50 | 50 | 50 |
| Treatment of malaria cases at health facilities (%) | 38.10 | 38 | 38 | 38 |

## 2 Results

The metapopulation model is fitted to estimated yearly incidence rate and prevalence among children aged two to ten years old from Malaria Atlas Project from 2010 to 2018 presented in Figures 4 and 5, and then cross-validated against the estimates from 2019 to 2022. The model is run deterministically from 1990 to reach a steady state before being fitted to data from 2010. The fitting method used is the maximum likelihood approach by assuming an underlying Poisson distribution with rate *λ* as the number of cases seeking treatment per year. Several parameters are estimated through the data-fitting process using the L-BGFS optimisation algorithm. The model with the estimated parameter values is then run for a further four years and compared to data between 2019 and 2022.

### 2.1 Model fitting and validation

The Figures 9 and 10 show simulated (red for fitting, blue for validation) and observed malaria incidence (estimate of reported treated cases per 1000) (black) in Atacora, Borgou, Donga and Alibori from 2010 to 2022. Confidence intervals (red for fitting, blue for validation) illustrate the uncertainty in predictions and model accuracy during both periods. Confidence intervals are shown as shaded areas, with red for the fitting period and blue for validation, highlighting model performance.

**Fig. 9:**
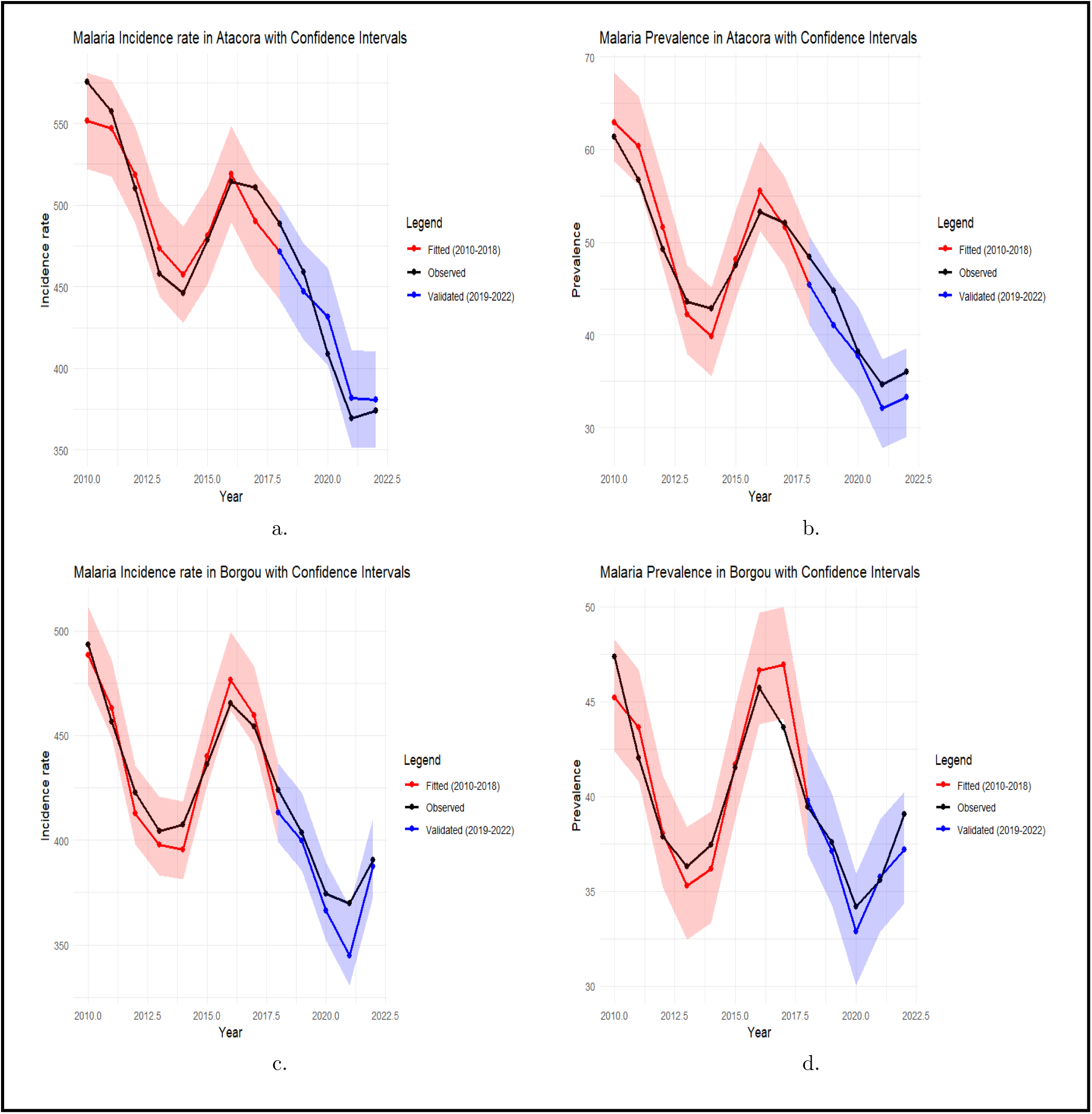
Data fitting and validation results for Atacora (a, b) and Borgou (c, d) regions in the northern part of Benin. The 10% uncertainty range for yearly incidence rate and prevalence predictions is shown.

**Fig. 10:**
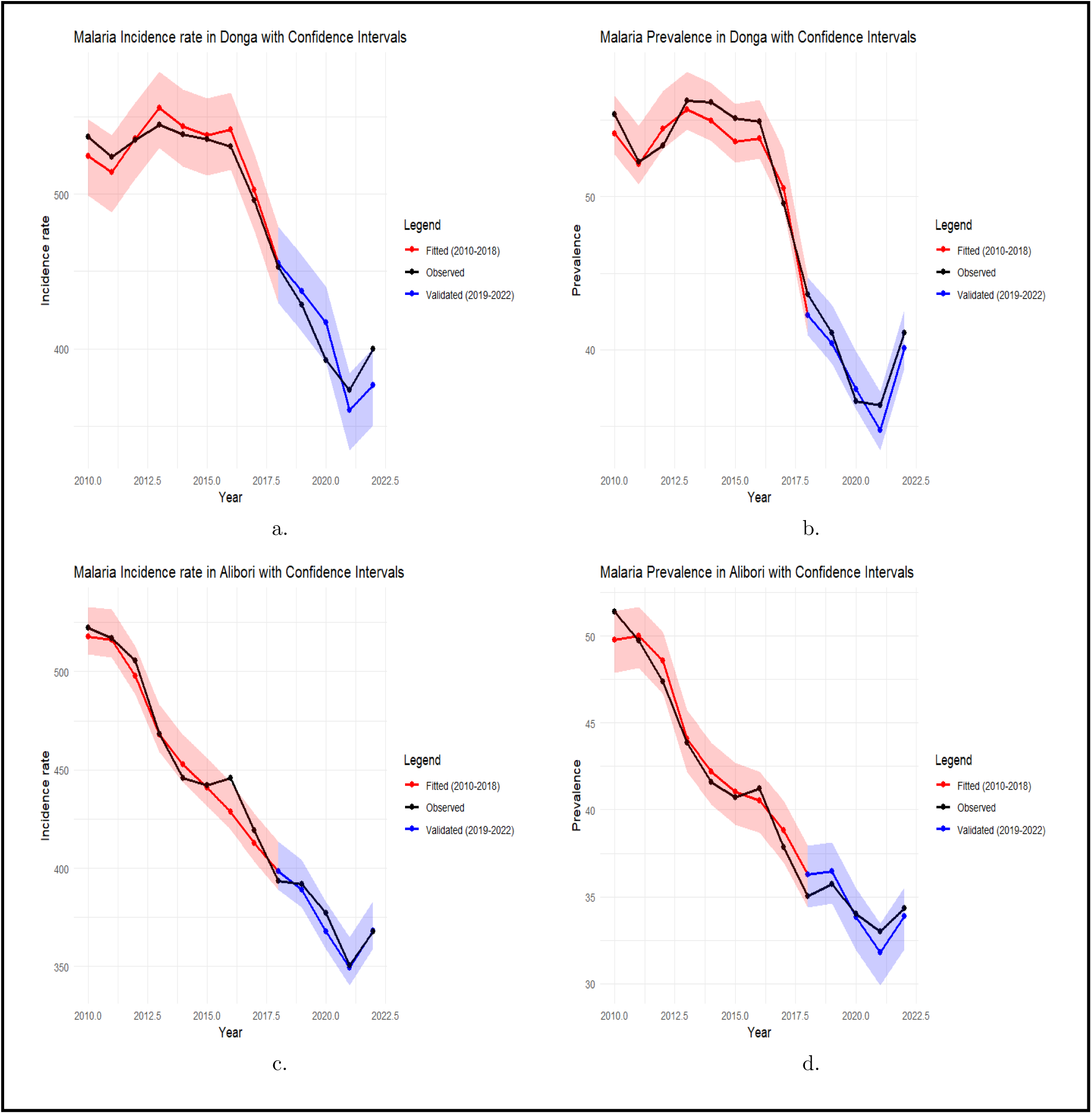
Data fitting and validation results for Donga (a, b) and Alibori (c, d)regions in the northern part of Benin. The 10% uncertainty range for yearly incidence rate and prevalence predictions is shown.

The model fitting results demonstrated a strong agreement with the estimates incidence (estimate of reported treated cases per 1000) and estimate prevalence from Malaria Atlas Project across the four studied regions: Atacora, Alibori, Donga, and Borgou. The Poisson likelihood approach, combined with the L-BFGS optimization algorithm, successfully estimated key parameters, ensuring that the model closely matched historical malaria burden data. Figures 9 and 10 illustrate the fitted and predicted incidence rates with their respective 95% confidence intervals. The parameters obtained from the data fitting process are provided in the Supplementary file.

The validation process further confirmed the robustness of the model, as predictions for the post-fitting period (2019-2022) remained within the expected confidence intervals. This suggests that the model effectively captures the main drivers of malaria transmission and can be reliably used to assess intervention scenarios.

### 2.2 Combining interventions

To evaluate the potential impact of enhanced malaria control, we simulated various intervention scale-up scenarios and assessed their effects on malaria incidence in the northern region of Benin. The results, presented in Figures 11, 12, 13 and 14 illustrate the projected trends in incidence (estimate of clinical cases per 1000 among children under five and estimate of reported treated cases per 1000) rates under each scenario over a twenty-five-year time horizon.

**Fig. 11:**
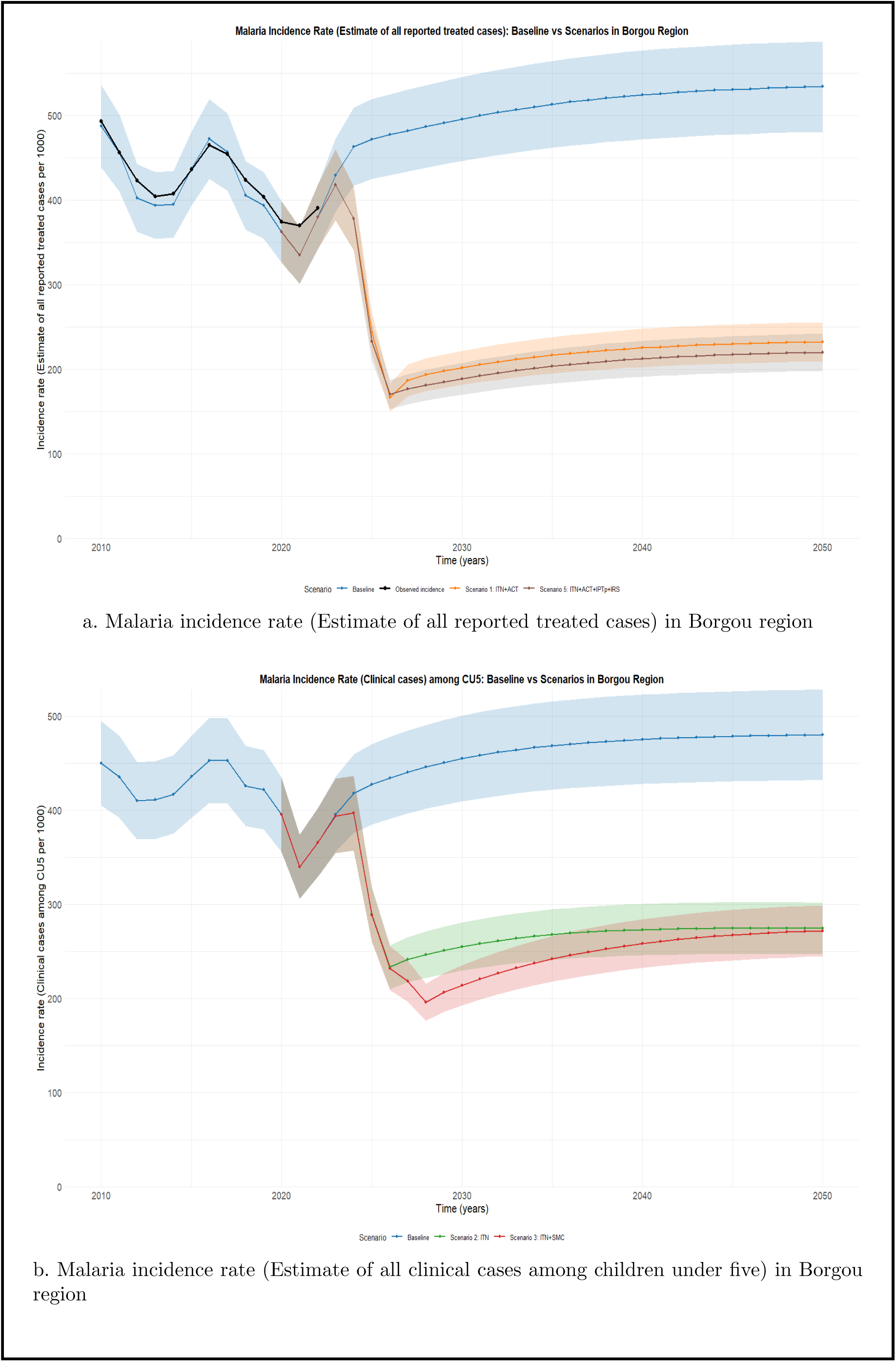
Predicted impact of interventions on incidence rate (estimate of clinical cases per 1000 among children under five and estimate of reported treated cases per 1000) in Borgou region

**Fig. 12:**
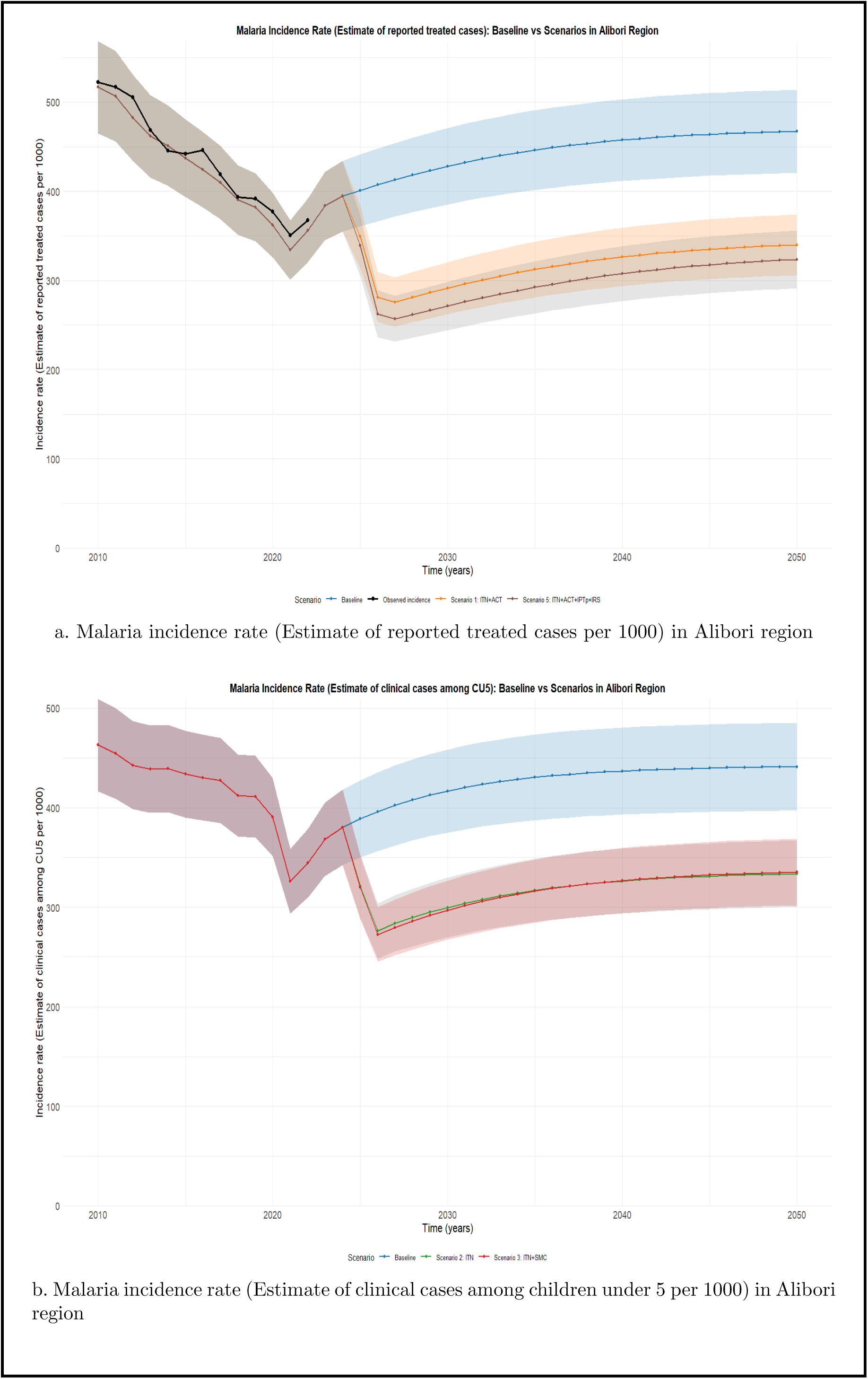
Predicted impact of interventions on incidence rate (estimate of clinical cases per 1000 among children under five and estimate of reported treated cases per 1000) in Alibori region

**Fig. 13:**
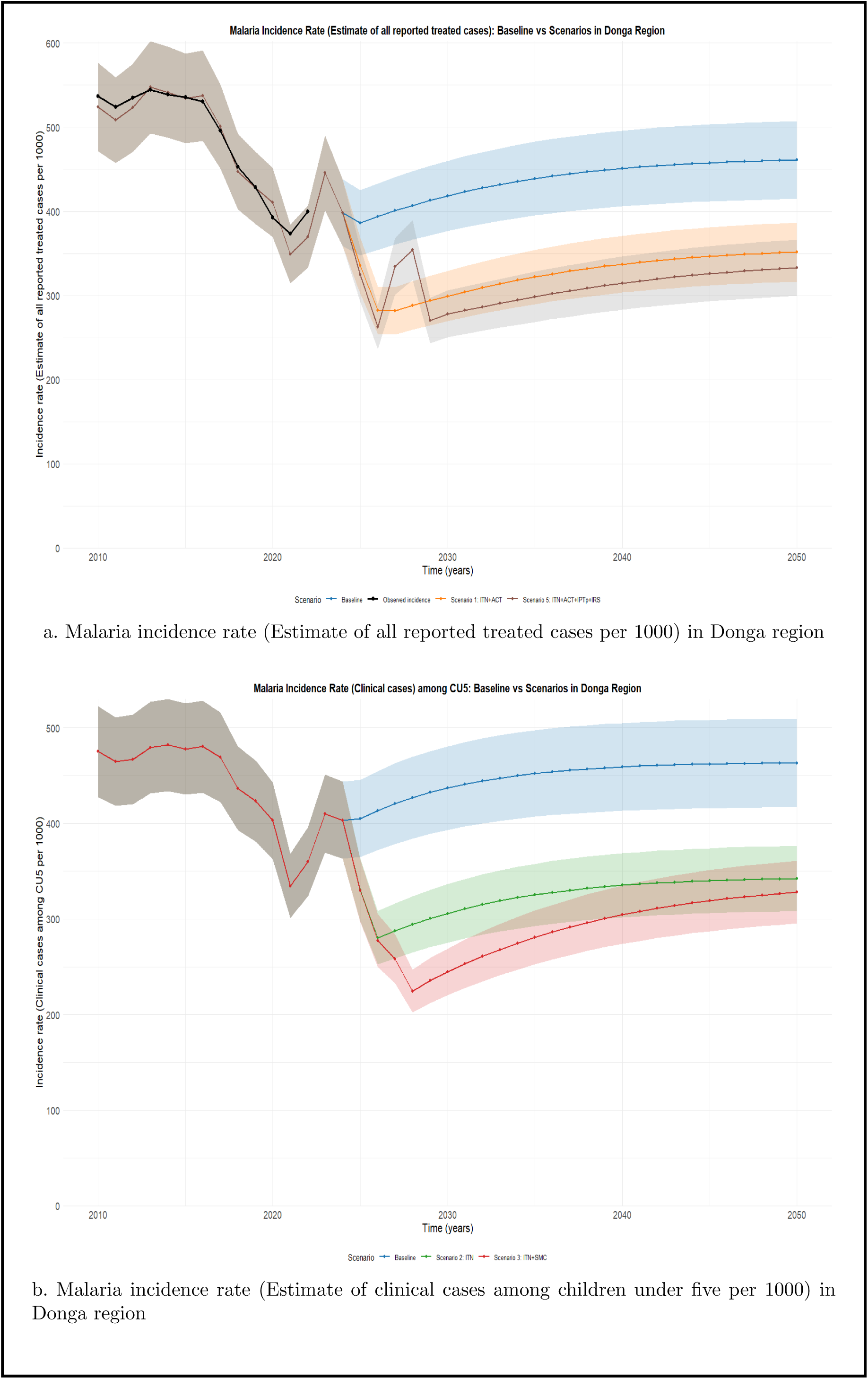
Predicted impact of interventions on incidence rate (estimate of clinical cases per 1000 among children under five and estimate of reported treated cases per 1000) in Donga region

**Fig. 14:**
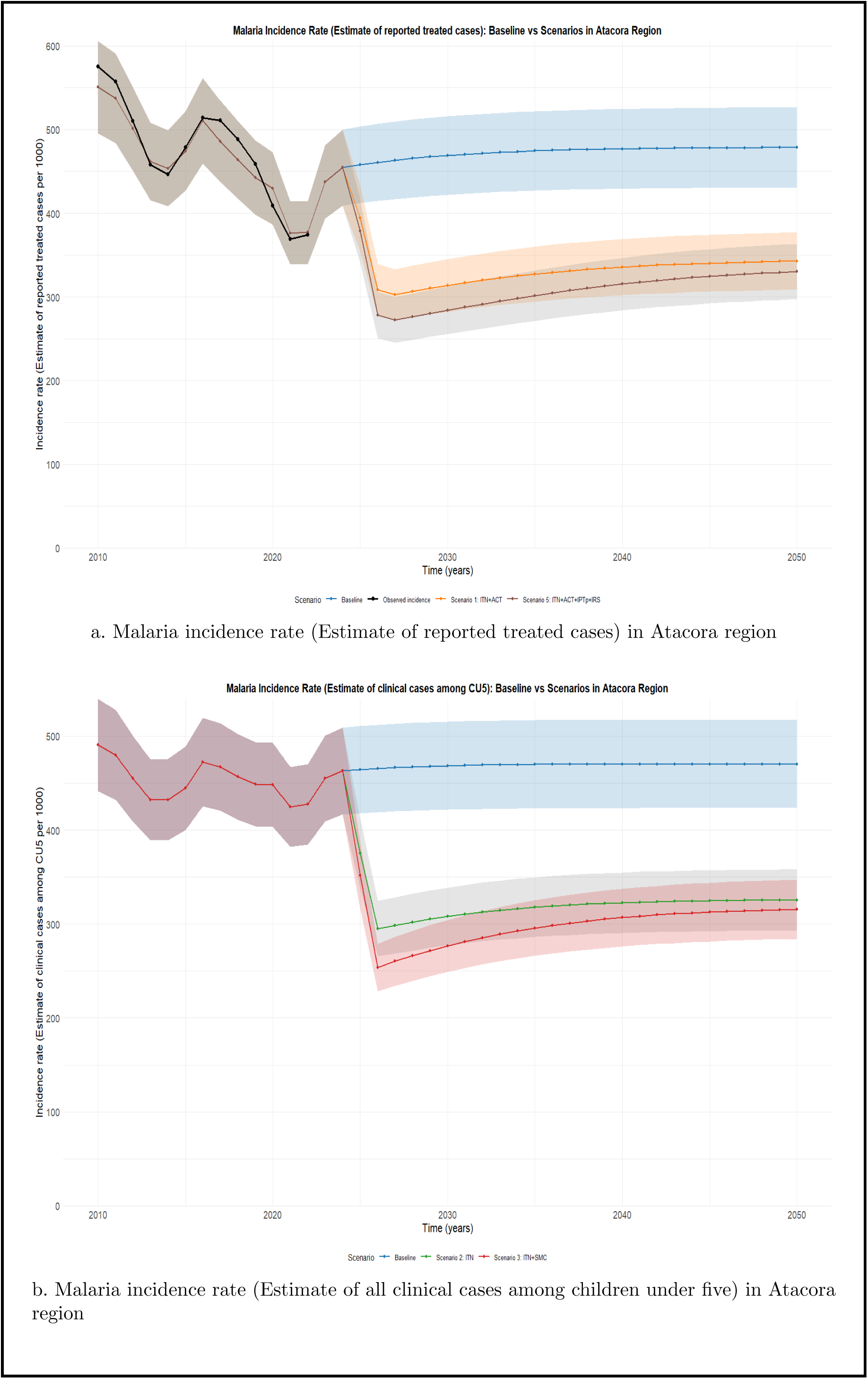
Predicted impact of interventions on incidence rate (estimate of clinical cases per 1000 among children under five and estimate of reported treated cases per 1000) in Atacora region

The figures 11, 12, 13 and 14 present the projected malaria incidence rates per 1000 (estimate of reported treated cases and estimates of clinical cases among children under five) in Atacora, Borgou, Donga and Alibori regions under various intervention scenarios from 2025 to 2050. The baseline scenario, representing the current intervention coverage, is compared to four alternative scenarios:

- Scaling up ITN (Insecticide-Treated Nets) coverage combined with effective ACT (Artemisinin-based Combination Therapy) treatment coverage.
- Scaling up ITN (Insecticide-Treated Nets) coverage alone
- Scaling up ITN coverage combined with SMC (Seasonal Malaria Chemoprevention) coverage.
- Scaling up ITN, SMC, IPTp, IRS, and effective antimalarial treatment.

#### 2.2.1 General Patterns and Seasonal Changes

When comparing the estimated incidence of reported treated malaria cases between Alibori and Borgou, both regions display similar historical trends and intervention impacts, but Borgou demonstrates slightly greater reductions under intensified strategies. Under baseline conditions, both regions show a rise in incidence (estimate of reported treated cases) after 2023, with Alibori stabilizing near 470 cases per 1000 and Borgou approaching 530 per 1000 by 2050. Scenario 1 (ITN + ACT) reduces incidence in both regions, but the impact is more pronounced in Borgou. Scenario 5 (ITN + ACT + IPTp + IRS) yields the most substantial reductions, bringing incidence down to just under 330 per 1000 in Alibori and approximately 310 per 1000 in Borgou by 2050.

The estimated incidence of clinical malaria cases among children under five (CU5) reveals important contrasts between Alibori and Borgou, both in baseline burden and intervention effectiveness. Under the baseline scenario, incidence in both regions initially declines but begins to rise again post-2023, reaching approximately 440 cases per 1000 in Alibori and slightly higher in Borgou, nearing 470 per 1000 by 2050. Scenario 2 (ITN alone) results in moderate reductions in both regions; however, the incidence remains relatively high, stabilizing around 350-360 per 1000 in both settings. Notably, the addition of Seasonal Malaria Chemoprevention (SMC) in Scenario 3 significantly enhances impact, particularly in Borgou, where incidence is projected to decline to just above 260 per 1000, compared to about 340 per 1000 in Alibori. This suggests that Borgou’s transmission profile and health system may be more responsive to SMC, potentially due to higher transmission seasonality or more favorable demographic characteristics. The sharper decline observed in Borgou also reflects greater potential for accelerated control in this region, provided that high coverage and adherence to SMC and ITN use can be maintained.

The projected trends in the incidence of reported treated malaria cases in the Atacora region demonstrate the significant potential impact of intensified malaria control interventions. Historically, the incidence followed a fluctuating but generally decreasing trend from 2010 to 2022, with observed and modeled estimates closely aligned, stabilizing around 380-400 cases per 1000 by 2022. Under the baseline scenario, which assumes no additional scale-up of interventions, incidence gradually increases after 2023, reaching nearly 480 cases per 1000 by 2050. This rebound highlights the limitations of current control efforts and the likelihood of resurgence without intensified action.

In Scenario 1 (ITN + ACT), a notable reduction in incidence (estimate of reported treated cases per 1000) is observed, with levels stabilizing around 370 cases per 1000 by 2050. This suggests that improved case management (ACT) and vector control (ITNs) alone can yield moderate reductions. However, Scenario 5, which combines ITN, ACT, IPTp, and IRS, offers the most substantial improvement. The incidence under this integrated strategy is projected to drop to about 340 cases per 1000 by 2050, marking a sustained and more pronounced decline compared to the other scenarios.

The incidence of reported treated malaria cases in Donga reveals a more volatile historical pattern compared to other northern regions. Between 2010 and 2022, the incidence fluctuates substantially, peaking above 560 cases per 1000 around 2015 and again around 2019, before declining sharply to below 400 per 1000 by 2022. This high variability suggests a region with potentially unstable transmission dynamics, inconsistent intervention coverage, or reporting challenges. Under the baseline scenario, incidence (estimate of reported treated cases per 1000) increases post-2023, reaching about 470 per 1000 by 2050, indicating a significant risk of rebound in the absence of intervention intensification. Scenario 1 (ITN + ACT) lowers this burden moderately, stabilizing at around 390 per 1000. However, the most notable reduction is observed in Scenario 5 (ITN + ACT + IPTp + IRS), where incidence falls to approximately 360 per 1000 by 2050. Despite these improvements, Donga’s projections remain higher than those of most other regions under the same scenarios, reflecting persistent transmission risk and perhaps greater challenges in achieving sustained control.

#### Cross-Regional Observations

The comparative analysis of projected malaria incidence in terms of reported treated cases across the four northern regions of Benin such as Alibori, Atacora, Borgou, and Donga reveals that while all regions would benefit from intensified intervention strategies, the scale of impact and responsiveness varies. Under the baseline scenario, all regions exhibit a gradual resurgence in incidence beyond 2023, reflecting the limitations of maintaining current control levels. Scenario 1 (ITN + ACT) yields moderate but consistent reductions, whereas Scenario 5 (ITN + ACT + IPTp + IRS) produces the most substantial and sustained declines across all regions. Borgou shows the most favorable response, with projected incidence falling below 300 per 1000, followed by Atacora, which maintains a relatively low burden under both scenarios. In contrast, Alibori and Donga present higher projected burdens even under Scenario 5, suggesting potential implementation challenges or higher baseline transmission. Importantly, none of the regions reach pre-elimination thresholds, highlighting that while integrated strategies offer measurable gains, achieving malaria elimination in these regions may require additional innovations, sustained coverage, and region-specific adaptations. These findings underscore the critical need for tailored, comprehensive, and well-implemented intervention packages to accelerate malaria control and progress toward elimination in northern Benin.

The analysis of projected clinical malaria incidence among children under five reveals regional differences in both baseline burden and the expected impact of intervention strategies. Across all four regions, the baseline scenario (no intensified interventions) shows a resurgence in incidence after 2023, stabilizing around 420-470 cases per 1000 by 2050. This pattern underscores the risk of rebound in the absence of enhanced control measures. The ITN-only scenario (Scenario 2) leads to moderate improvements across regions, with incidence dropping by approximately 60-100 cases per 1000, but remaining far from elimination thresholds.

The greatest gains are achieved under Scenario 3 (ITN + SMC), which consistently results in the lowest projected incidence among CU5. Notably, Borgou shows the most substantial decline under this combined strategy, with incidence falling to just above 260 per 1000, suggesting high responsiveness to SMC. Atacora also demonstrates significant improvement, reaching close to 300 per 1000, while Alibori and Donga stabilize at higher levels (around 330-340 per 1000), indicating a relatively smaller marginal benefit from SMC in those regions.

## Discussion

In partnership with funders, the Republic of Benin has developed subnationally tailored intervention plans for the National Malaria Control Program (NMCP). While several scholars [9, 11–17] have contributed to understanding malaria dynamics in Benin, this study represents the first application of mathematical modeling to support subnational strategy development in the country. The potential effects of malaria interventions in Benin Republic were assessed by simulating different implementation scenarios. As said earlier, Benin has adopted ITN (Insecticide-Treated Nets) which are bed nets treated with insecticide that protect individuals from mosquito bites and kill mosquitoes, especially during sleeping hours; SMC (Seasonal Malaria Chemoprevention) which is Intermittent administration of antimalarial medicine to children from July to October every year to prevent malaria; IRS (Indoor Residual Spraying) which is the application of insecticide on the walls and surfaces inside homes to kill mosquitoes that rest indoors; IPTp (Intermittent Preventive Treatment in Pregnancy) which are scheduled administration of antimalarial drugs to pregnant women to prevent malaria infection during pregnancy and finally Anti-malarial Treatment which are effective medications, such as artemisinin-based combination therapies (ACTs), to treat individuals diagnosed with malaria. The models incorporate all these interventions and population sizes across the northern regions of Benin Republic.

In the litterature, the impact of combining interventions at a certain coverage to assess malaria elimination has been demonstrated by many scholars [18, 20, 21, 37]. The results of this study now underscore the critical role of fully integrated malaria interventions, particularly Scenario 5 (ITNs + IRS + Treatment + SMC + IPTp), in controlling malaria incidence across all four regions. The large decrease in cases seen under Scenario 5 highlights the importance of mixing preventive measures such as insecticide-treated nets (ITNs) [20, 21] and indoor residual spraying (IRS) [18] with effective treatment and other protective strategies (SMC and IPTp). This layered approach reduces human vector contact and ensures timely treatment of cases, breaking the transmission cycle at many points. The steady and rapid decline in malaria incidence under this scenario shows its potential for achieving elimination in these four regions of the northern part of Benin Republic.

Looking at the current situation in Benin, Long-Lasting Insecticidal Nets (LLINs) are the most used for malaria prevention. The National Malaria Control Programme (NMCP) has run several mass distribution campaigns to reach universal coverage [7]. However, ITN/LLIN usage is still low in many areas particularly in Atacora, Borgou, Alibori and Donga [40]. Our findings suggest that a decrease in malaria incidence may be possible with increased LLINs coverage and usage to 80% before 2028 in the northern regions where transmission is very high.

Scenarios 1, 2 and 3 also showed notable reductions in malaria incidence both in estimate of reported treated cases and clinical cases among children under five years old.

Interestingly, Scenario 1, which combines insecticide-treated nets (ITNs) with effective case management using antimalarial treatment (ACT), demonstrated a more substantial and sustained reduction in malaria incidence compared to the baseline and emerged as the second most effective strategy, significantly reducing malaria cases, though it was less impactful than the comprehensive approach involving all five interventions. This suggests that strengthening the core pillars of malaria control prevention through ITNs and timely, effective treatment may yield more consistent impact than a broader yet potentially harder-to-implement mix of interventions. For the NMCP of Benin, this result highlights the critical importance of reinforcing ITN distribution as said earlier and ensuring universal access to quality malaria diagnosis and treatment services. While integrated packages remain ideal in theory, their success is highly dependent on operational feasibility, sustained funding, and high community uptake. Scenario 1, being simpler and more cost-effective to implement, may offer a more practical and scalable strategy, particularly in resource-constrained settings. We recommend prioritizing and maintaining high ITN coverage and usage, investing in community health systems to ensure prompt diagnosis and treatment, and monitor stock levels to avoid treatment shortages. Additionally, efforts should be made to improve health-seeking behavior through community education and outreach, ensuring that people with fever access care promptly. These foundational interventions, if well-executed, can serve as a strong backbone for malaria control and potentially pave the way for elimination in these four regions of the northern part of Benin.

Scenarios 3 (ITN + SMC) showed similar trends in cutting incidence. These interventions, which target children under five (SMC), help lower malaria cases. The results of this study encourage to invest more in providing robust Seasonal Malaria Chemoprevention (SMC). Increasing investment in this intervention can help protect the most vulnerable groups which are young children from malaria, reduce treatment costs, and ultimately improve overall community health and economic development. Scaling (IRS) which coverage was maintained at 50% coverage had a lower effect on reducing the burden of malaria in the four regions. This is likely because IRS depends on very high coverage over time and faces issues like operational challenges and insecticide resistance.

In summary, the findings from this study highlight several important points for malaria control and elimination in Benin and show it looks to take a long time to eliminate malaria in the northern regions of Benin Republic. First, the success of Scenario 5 shows that using a full package of interventions addressing both prevention and case management is key. Standalone or partly integrated strategies, while they can lower incidence, may not be enough to completely control malaria, especially in these areas in the northern Benin with high and seasonal transmission.

Second, sustained resources and strong political commitment are needed to keep up an integrated strategy like Scenario 5. Controlling malaria incidence with ITNs, IRS, Treatment, SMC, and IPTp requires high community involvement, regular net distribution, ongoing health campaigns, and a robust health system to cover treatments.

Despite the strengths and comprehensive nature of this study, several limitations should be noted. The metapopulation model relies on several simplifying assumptions, such as full protection during SMC and IPTp treatment. These assumptions may not fully capture the complexities of real-world malaria transmission dynamics. In addition, the model calibration and validation depend on estimates data from the Malaria Atlas Project. The predicted estimates from the Malaria Atlas Project carry a high degree of uncertainty and should not be interpreted as precise forecasts. Any inaccuracies or gaps in these data sources can affect the precision of model predictions. Although several parameters were estimated through a maximum likelihood approach, inherent uncertainties in these values persist. This uncertainty is partially addressed through the inclusion of confidence intervals in the results (10%), but it remains a challenge for accurately predicting long-term outcomes. Furthermore, the model assumes that intervention coverage can be increased as specified in the scenarios. In reality, scaling up interventions such as IRS, ITN distribution, and effective case management faces operational, logistical, and behavioral challenges that are not fully captured in the simulations. In addition, there are some other limitations such as uncertain SMC and IPTp coverage in the four regions in the northern Benin, which may affect the accuracy of the intervention impact estimates and introduce variability in model projections. The model did not account for vaccination, which has been introduced since January 2024, and this omission may lead to an underestimation of future reductions in malaria transmission. Moreover, the model did not account for long-term trends in economic development, housing improvements, or environmental management, which may well reduce malaria risk for many Beninese over the next decade. Lastly, the model did not include climate change or year-to-year variations in climate. Addressing these limitations in future research could involve incorporating socioeconomic and environmental data into transmission models, integrating climate projections to capture potential variability in malaria risk, and exploring the combined effects of structural and behavioral changes on disease dynamics, incorporating more dynamic data sources from the NMCP of Benin Republic, refining model assumptions, including climate factors into the model and exploring additional intervention scenarios that account for real-world operational challenges.

Despite its limitations, the model successfully provided a quantitative assessment of the impact of a combination of interventions on malaria control in Benin over the next decade. In light of the model’s limitations, this project underscores the need for increased investment in the collection of subnational surveillance data in the Republic of Benin, as well as in research aimed at improving the understanding of spatial variation in malaria transmission, intervention coverage and effectiveness, and entomological factors. High-quality data is essential for evidence-based and appropriately targeted national plans, as modeling cannot substitute for inadequate or incomplete data.

Future research should then explore the cost-effectiveness and practical feasibility of implementing these comprehensive strategies not only in the four northern regions of the Benin Republic, but also in the eight remaining regions that were not covered in this study. Studies should also look at the impact of climate change on malaria transmission patterns, including how shifting temperatures and rainfall variability may influence vector dynamics, seasonality, and the geographic distribution of risk.

## 3 Conclusion

Mathematical modeling offers a framework for quantitatively assessing the potential impact of intervention scenarios. This study shows that a fully integrated approach using multiple interventions such as ITNs, IRS, Treatment, SMC, and IPTp is the most effective strategy for reducing malaria incidence and moving toward elimination in the northern regions of Benin. While each intervention alone or in smaller combinations can lower case numbers, only the comprehensive package (Scenario 5) achieved a rapid decline in malaria cases. The package (ITN + Treatment) also achieved a decline in malaria cases in Borgou, Donga, Atacora and Alibori and should not be neglected if we want to work towards malaria elimination in those regions. Lastly, there is an urgent need for investment in subnational data on malaria burden in Benin Republic and intervention effects to enable data-driven impact modeling at a more detailed level and to support better decision-making on optimal malaria intervention strategies at the subnational scale.

## Supporting information

Supplementary file 1: Model description

## Data Availability

All data used in the present work are available from public sources that are are contained in the manuscript

## Acknowledgments

We are grateful to the Bill and Melinda Gates foundation (INV 047-048) through the Modelling and Simulation Hub Africa for financial support.

## Author Contributions

**Conceptualization:** Kounoumi Sara Medekon, Sheetal Silal.

**Formal analysis:** Kounoumi Sara Medekon.

**Funding acquisition:** Sheetal Silal.

**Investigation:** Kounoumi Sara Medekon. **Methodology:** Kounoumi Sara Medekon, Sheetal Silal. **Resources:** Sheetal Silal.

**Software:** Kounoumi Sara Medekon.

**Supervision:** Sheetal Silal.

**Validation:** Sheetal Silal.

**Visualization:** Kounoumi Sara Medekon.

**Writing – original draft:** Kounoumi Sara Medekon.

**Writing – review & editing:** Jules Degila, Sheetal Silal.

