## Supplementary file 1: Model description for "Exploring pathways to malaria control and elimination in the northern regions (Atacora, Alibori, Borgou, Donga) of Benin Republic: a metapopulation modelling approach"


This appendix provides supplementary material to support the main text. It includes a detailed description of the model transmission dynamics for children, adults, and pregnant women. We also present the mathematical formulation used to assess malaria elimination in the northern regions of Benin, along with key assumptions and the expression for the force of infection. Furthermore, we summarize relevant literature on malaria modelling in Benin to give context to our work. The complete set of model equations, parameter values used across regions, and the definitions of all compartments in the model are also provided for transparency and reproducibility.

### Appendix A Model of transmission

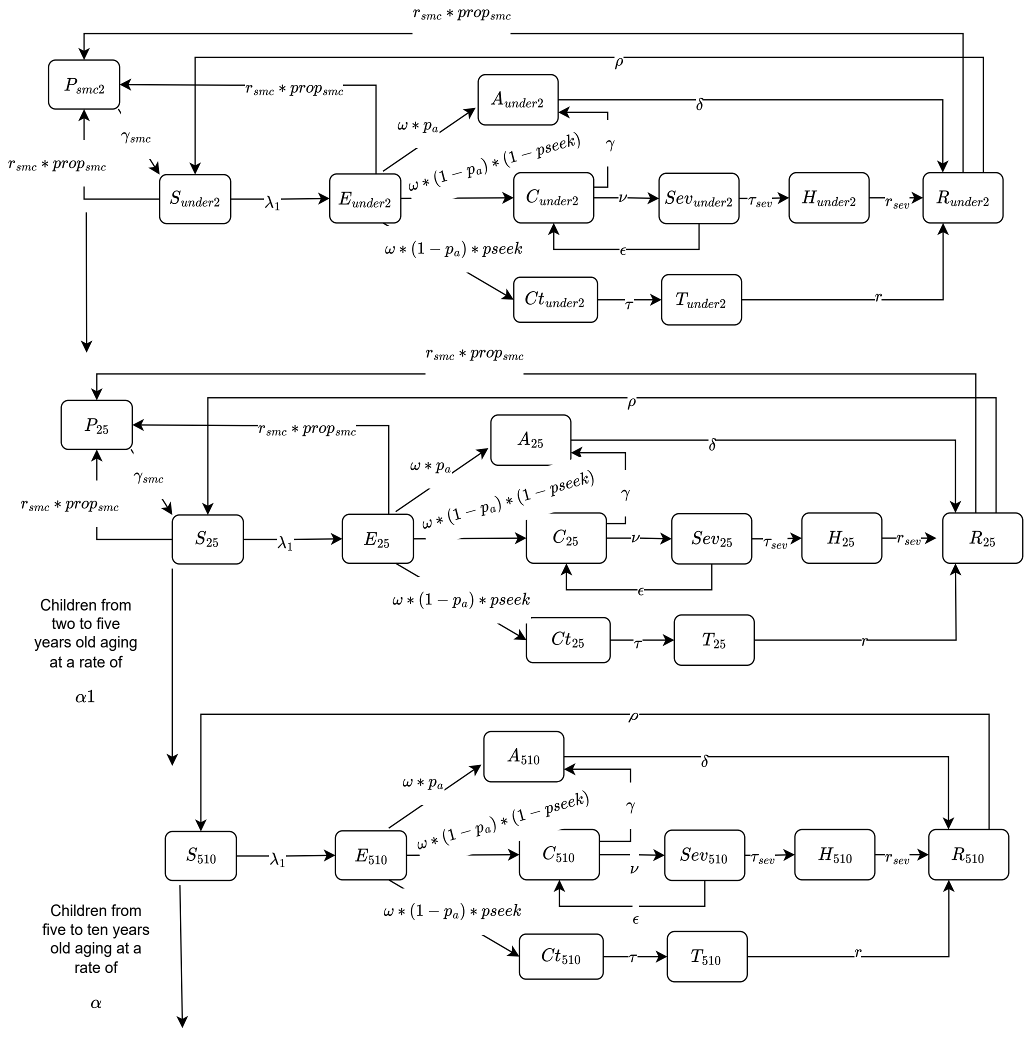


**Fig. A1**: Model of transmission for children

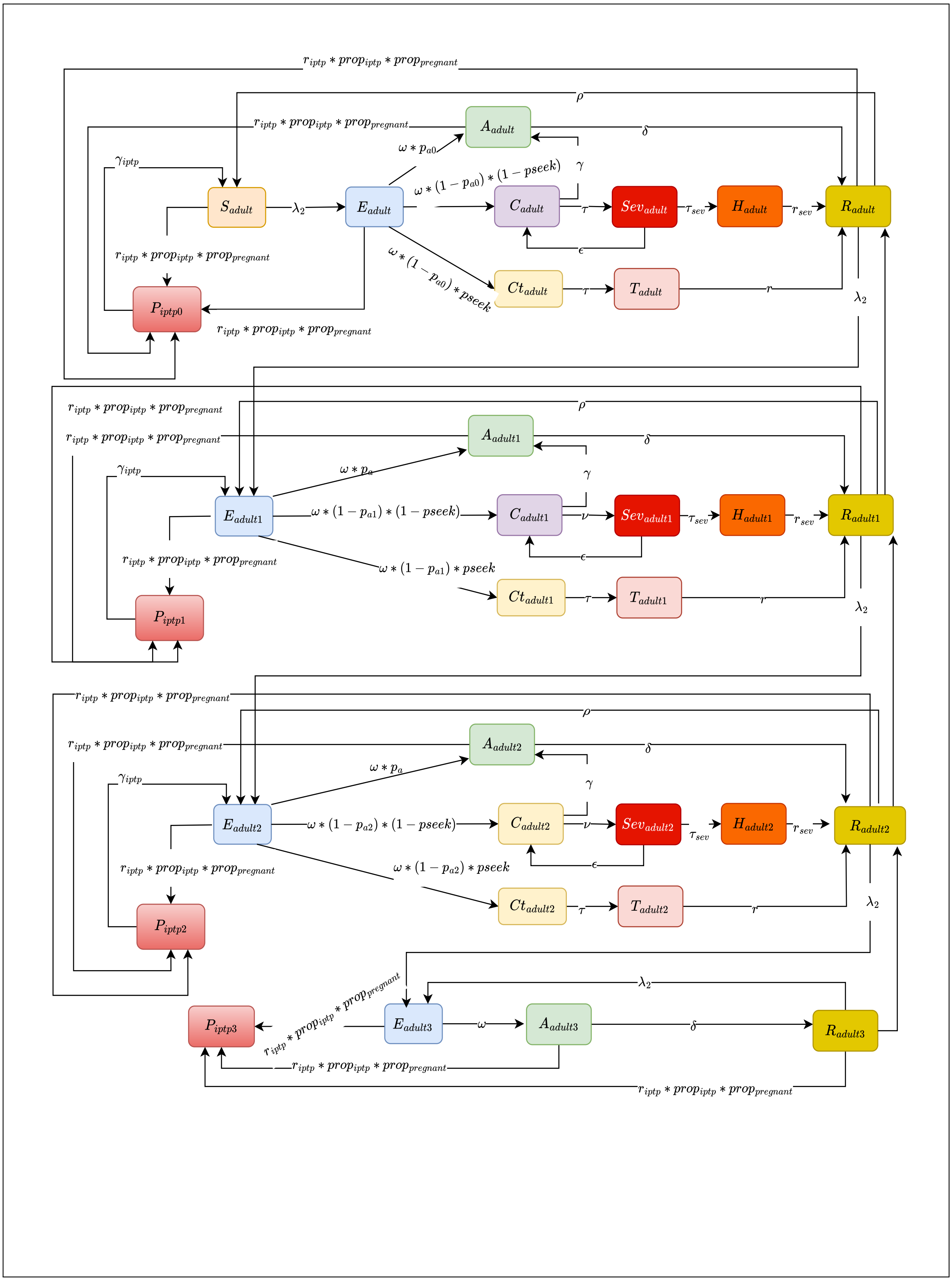


**Fig. A2**: Model of transmission for adults and pregnant women with the three levels of immunity

### Appendix B Assessing elimination of malaria

In the model, elimination is defined as achieving and maintaining zero locally acquired malaria cases in a specific region for three consecutive years, in line with the WHO's definition [1, 2]. As illustrated in Figure B3, the pathway to elimination typically starts with a control phase, where the goal is to reduce the Slide Positivity Rate (SPR) to below 5 per 1000. This is followed by a pre-elimination phase, targeting an SPR of less than 1 per 1000. Once sustained zero local transmission is achieved over three years, the region is considered to have eliminated malaria.

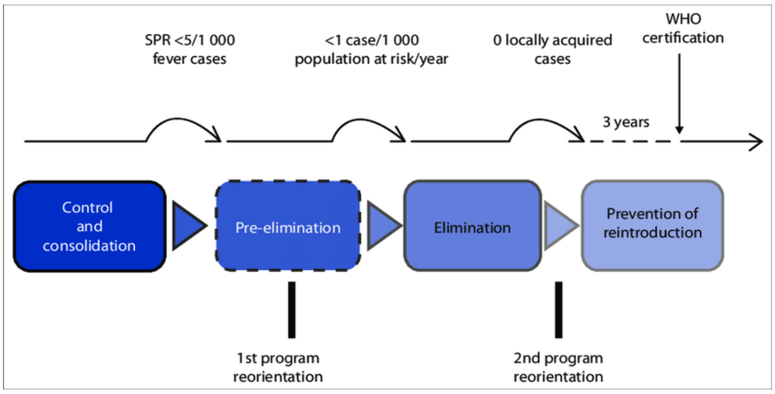


**Fig. B3**: Elimination process of malaria[3]

##### B.0.1 Additional Assumptions:

- The population is stratified into age groups with age-specific parameters.
- IPTp is given to all pregnant women without testing if they are infected with the malaria parasite;
- IPTp is not administered concurrently with an other anti-malaria drug;
- Children with active malaria infection do not recieve SMC treatment;
- Children are not getting malaria infection during prophylaxis of SMC treatment. They are fully protected;
- Pregnant women are not getting malaria infection during prophylaxis of IPTp treatment. They are fully protected
- Birth and death rates are not constant, and migration is not considered.

##### B.0.2 Seasonality

Seasonality is a critical aspect of malaria transmission, particularly in regions like Benin, where mosquito populations and biting rates fluctuate throughout the year due to climatic factors such as rainfall, temperature, and humidity. To accurately capture these seasonal dynamics, the model incorporates a seasonal forcing function that modulates the transmission rate over time. These seasonal variations are crucial for evaluating the timing and effectiveness of interventions, such as seasonal malaria chemoprevention (SMC), which targets high-transmission periods.

$$\text{seas} = 1 + \text{amp}_1 \cdot \left(\cos\left(\frac{2\pi \cdot \text{times}}{365} - \phi_1\right)\right)^{\text{peak}} + \text{amp}_2 \cdot \left(\cos\left(\frac{4\pi \cdot \text{times}}{365} - \phi_2\right)\right)^{\text{peak}} \tag{B1}$$

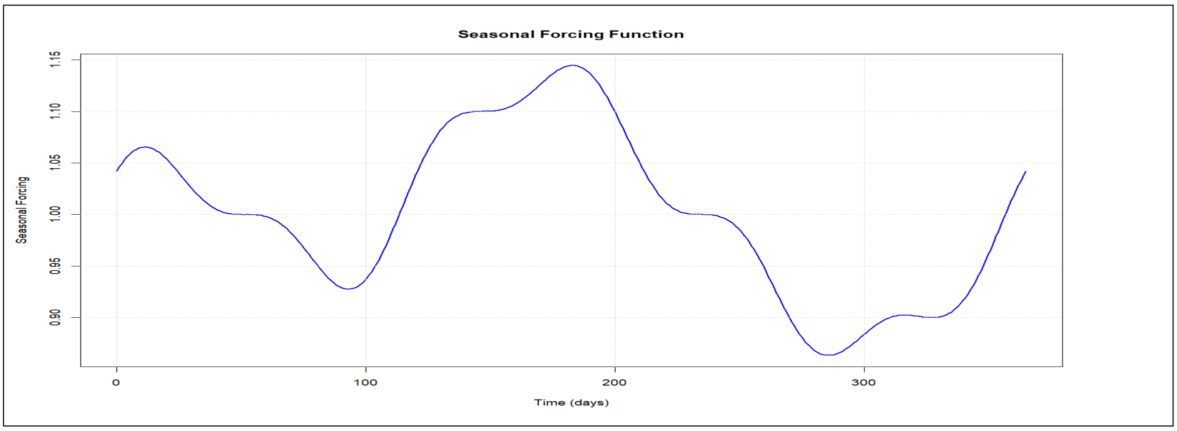


**Fig. B4**: Plot of the seasonal forcing function

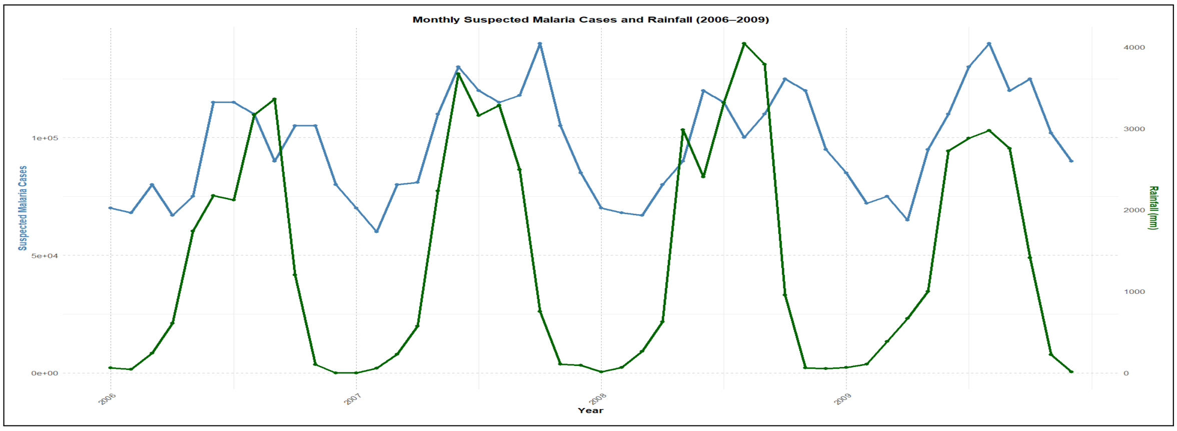


**Fig. B5**: Monthly malaria cases from 2006 to 2009 obtained from [4] used to determine the shape of the seasonal forcing function

The seasonal forcing function used in the model B4 was derived based on the observed relationship between monthly rainfall patterns and suspected malaria cases recorded between 2006 and 2009 B5. As rainfall strongly influences mosquito breeding and consequently malaria transmission, the temporal correlation between the two variables was analyzed. Peaks in rainfall consistently preceded or coincided with increases in malaria incidence, reflecting the typical lag between environmental conditions favorable for vector proliferation and the onset of human infections. Using this relationship, a smooth, periodic function was fitted to capture the seasonal variation in transmission intensity throughout the year. This function was incorporated into the model as a multiplicative seasonal factor to modulate the force of infection, thereby accounting for the cyclical nature of malaria transmission driven by climatic conditions.

##### B.0.3 Force of Infection

The force of infection, $\lambda_2$, represents the rate at which susceptible individuals acquire malaria infections within the population. It is influenced by various biological, environmental, and intervention-related factors, and in the model, it is defined as:

$$\lambda_2 = \text{seas} \cdot (1 - \text{itn} \cdot \text{itn\_eff} \cdot \text{itn\_usage\_2}) \cdot (1 - \text{irs} \cdot \text{irs\_eff}) \cdot \frac{a^2 bcm \frac{\text{Infectious}}{P}}{ac\frac{\text{Infectious}}{P} + \mu_m} \cdot \frac{\gamma_m}{\gamma_m + \mu_m}$$

### Appendix C Summary of Benin Articles on Malaria Modelling and Understanding Using Modelling

**Table C1**: Summary of Benin Articles on Malaria Modelling and Understanding Using Modelling

| Reference | Study Focus | Methods | Key Findings/Conclusions |
|---|---|---|---|
| Moiroux et al. [5] | Abundance of malaria vectors in 19 villages of the Ouidah-Kpomassè-Tori Bossito (OKT) health administrative region in southern Benin (on the Atlantic coast)southern Benin | Negative binomial zero-truncated mixed-effect models | Moiroux et al. [5] built predictive maps revealing distinct seasonal and spatial abundance patterns for An. funestus, An. coluzzii, and An. gambiae s.s. |
| Sovi et al. [6] | Resistance status of Anopheles gambiae s.l. to insecticides following the 2011 mass distribution campaign of long-lasting insecticidal nets (LLINs) in the Plateau Department, south-eastern Benin | WHO susceptibility tests combined with PCR-based molecular assays using logistic regression | Sovi et al. [6] found that after the 2011 LLIN distribution, mosquitoes became much less sensitive to pyrethroids and DDT, and the kdr L1014F mutation became more common. Their results show why strong plans are needed to manage insecticide resistance in Benin Republic. |

Continued on next page

**Table C1 – continued from previous page**

| Reference | Study Focus | Methods | Key Findings/Conclusions |
|---|---|---|---|
| Arab et al. [7] | Effects of weather/climate on malaria distribution | Hierarchical Bayesian modelling framework applied to annual malaria and climate data from ten countries in West Africa including Benin | Identified a significant negative association between malaria incidence and climate factors (average temperature and total precipitation) in Benin. |
| Camponovo et al. [8] | Impact of routine case management on malaria burden in Kenya, Mozambique and Benin | Stochastic agent-based transmission modelling using OpenMalaria | In their study, the model predicted that increasing effective treatment coverage—from a baseline of around 28% up to 60%—could substantially reduce malaria risk in Benin. Specifically, the model estimated that in high-risk areas (where the prevalence in children under five is above 0.3), about 39% of young children would transition to lower-risk categories when treatment coverage is increased. Additionally, in lower transmission settings, the prevalence among young children could drop by up to 50%, accompanied by a 35% reduction in overall incidence. These findings highlight the potential benefits of scaling up effective treatment in reducing the malaria burden in Benin. |
| Boussari et al. [9] | They modelled the seasonality of Anopheles An. coluzzii and An. gambiae biting rates in the South Benin | Latent class trajectory modelling with village-level random intercepts | Classified villages into distinct biting rate profiles correlated with rainfall, altitude, larval site density, and vegetation index; supports targeted vector control strategies. They found that the biting rate profiles showed a strong correlation with factors such as average rainfall, altitude, the typical number of larval sites, and the average normalized difference vegetation index. |
| Keating et al. [10] | Assessment of malaria diagnostic capacity | Observational assessments with statistical analyses (logistic regression) | Keating et al. [10] found that malaria tests in Benin are generally done well and give accurate results. However, not everyone is tested at health facilities, and some people who test negative still receive antimalarial drugs. |
| Moiroux et al. [11] | Risk of being bitten by malaria vectors in a vector control area in southern Benin | Binomial mixed-effects models applied to presence-absence data from human landing catches | Produced predictive risk maps with high accuracy (AUC $>$ 0.9 for An. funestus); highlighted seasonal variations and environmental determinants of human-vector contact in 19 villages in the southern Benin. |

Continued on next page

**Table C1 – continued from previous page**

| Reference | Study Focus | Methods | Key Findings/Conclusions |
|---|---|---|---|
| Boussari et al. [12] | Vector density assessment | Non-parametric mixture of Poisson (NPMP) regression model with random effects | In order to assess the impact of TLLIN(Targeted Long-Lasting Insecticidal Nets), ULLIN(Universal Long-Lasting Insecticidal Nets), ULLIN+CTPS(Carbamate-Treated Plastic Sheeting), and TLLIN+IRS, Boussari et al. [12] designed a study where villages in the southern of Benin were randomly assigned to one of these four intervention groups. They then compared key entomological indicators such as malaria vector density, human biting rates, and overall transmission risk across the different groups. This approach allowed them to evaluate whether targeted LLIN coverage (TLLIN) or universal LLIN coverage (ULLIN) was more effective, and how the addition of complementary measures (either IRS or CTPS) further enhanced the control of malaria transmission. They found that vector density was not significantly different between treatment arms (TLLIN, ULLIN, TLLIN+IRS, and ULLIN+CTPS). The model helped to handle overdispersion and excess zeros in mosquito count data; showed that proximity to water bodies, market gardening, and high rainfall are linked to higher vector density, while water conveyance and cattle breeding are linked to lower density. |
| Cottrell et al. [13] | Influence of local environmental factors on transmission in the southern Benin | Three-level Poisson mixed regression model | Demonstrated that local climatic and household environmental factors (e.g., soil type, vegetation index, proximity to watercourses) explain significant spatial and temporal variations in vector density, enabling predictive mapping of transmission risk. |

### Appendix D Model equations

#### Equations

The full set of equations is presented below, followed by a table summarizing the model parameters, their values used for each region (Atacora, Alibori, Donga, and Borgou), and their corresponding sources. A detailed table of all model compartments and their definitions is also included to provide clarity on the model structure.

#### Equations

##### Children under 2

$$\frac{dS_{\text{under2}}}{dt} = \mu_b P - \lambda_1 S_{\text{under2}} + \gamma_{\text{smc}} P_{\text{smc2}} - r_{\text{smc}} \cdot \text{seasonal}_{\text{smc}} S_{\text{under2}} + \rho R_{\text{under2}} - \mu_h S_{\text{under2}} - \alpha_2 S_{\text{under2}}$$

$$\frac{dE_{\text{under2}}}{dt} = \lambda_1 S_{\text{under2}} - \omega E_{\text{under2}} - \alpha_2 E_{\text{under2}} - \mu_h E_{\text{under2}} - r_{\text{smc}} \cdot \text{seasonal}_{\text{smc}} E_{\text{under2}}$$

$$\frac{dA_{\text{under2}}}{dt} = \omega p_a E_{\text{under2}} - \delta A_{\text{under2}} + \gamma C_{\text{under2}} - \alpha_2 A_{\text{under2}} - \mu_h A_{\text{under2}}$$

$$\frac{dC_{\text{under2}}}{dt} = \omega(1-p_a)(1-p_{\text{seek}}) E_{\text{under2}} - \gamma C_{\text{under2}} - \alpha_2 C_{\text{under2}} - \mu_h C_{\text{under2}} - \nu C_{\text{under2}} + \epsilon \text{Sev}_{\text{under2}}$$

$$\frac{dCt_{\text{under2}}}{dt} = \omega(1-p_a) p_{\text{seek}} E_{\text{under2}} - \tau Ct_{\text{under2}} - \alpha_2 Ct_{\text{under2}} - \mu_h Ct_{\text{under2}}$$

$$\frac{d\text{Sev}_{\text{under2}}}{dt} = \nu C_{\text{under2}} - (\epsilon + \tau_{\text{sev}} + \mu_{\text{sev1}} + \mu_h) \text{Sev}_{\text{under2}} - \alpha_2 \text{Sev}_{\text{under2}}$$

$$\frac{dH_{\text{under2}}}{dt} = \tau_{\text{sev}} \text{Sev}_{\text{under2}} - (r_{\text{sev}} + \mu_{\text{sev2}} + \mu_h) H_{\text{under2}} - \alpha_2 H_{\text{under2}}$$

$$\frac{dT_{\text{under2}}}{dt} = \tau Ct_{\text{under2}} - r T_{\text{under2}} - \alpha_2 T_{\text{under2}} - \mu_h T_{\text{under2}}$$

$$\frac{dR_{\text{under2}}}{dt} = \delta A_{\text{under2}} + r T_{\text{under2}} + r_{\text{sev}} H_{\text{under2}} - \rho R_{\text{under2}} - r_{\text{smc}} \cdot \text{seasonal}_{\text{smc}} R_{\text{under2}} - \alpha_2 R_{\text{under2}}$$

$$-\mu_h R_{\text{under2}}$$

$$\frac{dP_{\text{smc2}}}{dt} = r_{\text{smc}} \cdot \text{seasonal}_{\text{smc}} S_{\text{under2}} - \gamma_{\text{smc}} P_{\text{smc2}} + r_{\text{smc}} \cdot \text{seasonal}_{\text{smc}} R_{\text{under2}} - \mu_h P_{\text{smc2}} + r_{\text{smc}} \cdot \text{seasonal}_{\text{smc}} E_{\text{under2}}$$

#### Children from 2 to 5

$$\frac{dS_{25}}{dt} = -\lambda_1 S_{25} - r_{\text{smc}} \cdot \text{seasonal}_{\text{smc}} S_{25} + \gamma_{\text{smc}} P_{\text{smc25}} + \rho_1 R_{25} - \mu_h S_{25} - \alpha_1 S_{25} + \alpha_2 S_{\text{under2}}$$

$$\frac{dE_{25}}{dt} = \lambda_1 S_{25} - \omega E_{25} - \alpha_1 E_{25} - \mu_h E_{25} - r_{\text{smc}} \cdot \text{seasonal}_{\text{smc}} E_{25} + \alpha_2 E_{\text{under2}}$$

$$\frac{dA_{25}}{dt} = \omega p_a E_{25} - \delta_1 A_{25} + \gamma C_{25} - \alpha_1 A_{25} - \mu_h A_{25} + \alpha_2 A_{\text{under2}}$$

$$\frac{dC_{25}}{dt} = \omega(1-p_a)(1-p_{\text{seek}}) E_{25} - \gamma C_{25} - \alpha_1 C_{25} - \mu_h C_{25} + \alpha_2 C_{\text{under2}} + \epsilon \text{Sev}_{25} - \nu C_{25}$$

$$\frac{dCt_{25}}{dt} = \omega(1-p_a) p_{\text{seek}} E_{25} - \tau Ct_{25} - \alpha_1 Ct_{25} - \mu_h Ct_{25} + \alpha_2 Ct_{\text{under2}}$$

$$\frac{d\text{Sev}_{25}}{dt} = \nu C_{25} - (\epsilon + \tau_{\text{sev}} + \mu_{\text{sev1}} + \mu_h) \text{Sev}_{25} - \alpha_1 \text{Sev}_{25} + \alpha_2 \text{Sev}_{\text{under2}}$$

$$\frac{dH_{25}}{dt} = \tau_{\text{sev}} \text{Sev}_{25} - (r_{\text{sev}} + \mu_{\text{sev2}} + \mu_h) H_{25} - \alpha_1 H_{25} + \alpha_2 H_{\text{under2}}$$

$$\frac{dT_{25}}{dt} = \tau Ct_{25} - r T_{25} - \alpha_1 T_{25} - \mu_h T_{25} + \alpha_2 T_{\text{under2}}$$

$$\frac{dR_{25}}{dt} = \delta_1 A_{25} + r T_{25} - \rho_1 R_{25} + r_{\text{sev}} H_{25} - r_{\text{smc}} \cdot \text{seasonal}_{\text{smc}} R_{25} - \alpha_1 R_{25} - \mu_h R_{25} + \alpha_2 R_{\text{under2}}$$

$$\frac{dP_{\text{smc25}}}{dt} = r_{\text{smc}} \cdot \text{seasonal}_{\text{smc}} S_{25} - \gamma_{\text{smc}} P_{\text{smc25}} + r_{\text{smc}} \cdot \text{seasonal}_{\text{smc}} R_{25} - \mu_h P_{\text{smc25}} + r_{\text{smc}} \cdot \text{seasonal}_{\text{smc}} E_{25}$$

#### Children from 5 to 10

$$\frac{dS_{510}}{dt} = -\lambda_1 S_{510} + \rho_2 R_{510} - \mu_h S_{510} - \alpha_3 S_{510} + \alpha_1 S_{25}$$

$$\frac{dE_{510}}{dt} = \lambda_1 S_{510} - \omega E_{510} - \alpha_3 E_{510} - \mu_h E_{510} + \alpha_1 E_{25}$$

$$\frac{dA_{510}}{dt} = \omega p_a E_{510} - \delta_2 A_{510} + \gamma C_{510} - \alpha_3 A_{510} - \mu_h A_{510} + \alpha_1 A_{25}$$

$$\frac{dC_{510}}{dt} = \omega(1-p_a)(1-p_{\text{seek}}) E_{510} - \gamma C_{510} - \alpha_3 C_{510} - \mu_h C_{510} + \alpha_1 C_{25} - \nu C_{510} + \epsilon \text{Sev}_{510}$$

$$\frac{dCt_{510}}{dt} = \omega(1-p_a) p_{\text{seek}} E_{510} - \tau Ct_{510} - \alpha_3 Ct_{510} - \mu_h Ct_{510} + \alpha_1 Ct_{25}$$

$$\frac{d\text{Sev}_{510}}{dt} = \nu C_{510} - (\epsilon + \tau_{\text{sev}} + \mu_{\text{sev1}} + \mu_h) \text{Sev}_{510} - \alpha_3 \text{Sev}_{510} + \alpha_1 \text{Sev}_{25}$$

$$\frac{dH_{510}}{dt} = \tau_{\text{sev}} \text{Sev}_{510} - (r_{\text{sev}} + \mu_{\text{sev2}} + \mu_h) H_{510} - \alpha_3 H_{510} + \alpha_1 H_{25}$$

$$\frac{dT_{510}}{dt} = \tau Ct_{510} - r T_{510} - \alpha_3 T_{510} - \mu_h T_{510} + \alpha_1 T_{25}$$

$$\frac{dR_{510}}{dt} = \delta_2 A_{510} + r T_{510} + r_{\text{sev}} H_{510} - \rho_2 R_{510} - \alpha_3 R_{510} - \mu_h R_{510} + \alpha_1 R_{25}$$

#### Adult with zero immunity Dynamics

$$\frac{dS_{\text{adult}}}{dt} = -\lambda_1 S_{\text{adult}} + \rho_3 R_{\text{adult}} - \mu_h S_{\text{adult}} - \alpha_4 S_{\text{adult}} + \alpha_3 S_{510}$$

$$\frac{dE_{\text{adult}}}{dt} = \lambda_1 S_{\text{adult}} - \omega E_{\text{adult}} - \alpha_4 E_{\text{adult}} - \mu_h E_{\text{adult}} + \alpha_3 E_{510}$$

$$\frac{dA_{\text{adult}}}{dt} = \omega p_a E_{\text{adult}} - \delta_3 A_{\text{adult}} + \gamma C_{\text{adult}} - \alpha_4 A_{\text{adult}} - \mu_h A_{\text{adult}} + \alpha_3 A_{510}$$

$$\frac{dC_{\text{adult}}}{dt} = \omega(1-p_a)(1-p_{\text{seek}}) E_{\text{adult}} - \gamma C_{\text{adult}} - \alpha_4 C_{\text{adult}} - \mu_h C_{\text{adult}} + \alpha_3 C_{510} - \nu C_{\text{adult}} + \epsilon \text{Sev}_{\text{adult}}$$

$$\frac{dCt_{\text{adult}}}{dt} = \omega(1-p_a) p_{\text{seek}} E_{\text{adult}} - \tau Ct_{\text{adult}} - \alpha_4 Ct_{\text{adult}} - \mu_h Ct_{\text{adult}} + \alpha_3 Ct_{510}$$

$$\frac{d\text{Sev}_{\text{adult}}}{dt} = \nu C_{\text{adult}} - (\epsilon + \tau_{\text{sev}} + \mu_{\text{sev1}} + \mu_h)\text{Sev}_{\text{adult}} - \alpha_4 \text{Sev}_{\text{adult}} + \alpha_3 \text{Sev}_{510}$$
$$\frac{dH_{\text{adult}}}{dt} = \tau_{\text{sev}}\text{Sev}_{\text{adult}} - (r_{\text{sev}} + \mu_{\text{sev2}} + \mu_h)H_{\text{adult}} - \alpha_4 H_{\text{adult}} + \alpha_3 H_{510}$$
$$\frac{dT_{\text{adult}}}{dt} = \tau Ct_{\text{adult}} - rT_{\text{adult}} - \alpha_4 T_{\text{adult}} - \mu_h T_{\text{adult}} + \alpha_3 T_{510}$$
$$\frac{dR_{\text{adult}}}{dt} = \delta_3 A_{\text{adult}} + rT_{\text{adult}} + r_{\text{sev}}H_{\text{adult}} - \rho_3 R_{\text{adult}} - \alpha_4 R_{\text{adult}} - \mu_h R_{\text{adult}} + \alpha_3 R_{510}$$
$$\frac{dS_{\text{adult1}}}{dt} = -\lambda_1 S_{\text{adult1}} + \rho_3 R_{\text{adult1}} - \mu_h S_{\text{adult1}}$$
$$\frac{dE_{\text{adult1}}}{dt} = \lambda_1 S_{\text{adult1}} - \omega E_{\text{adult1}} - \mu_h E_{\text{adult1}}$$
$$\frac{dA_{\text{adult1}}}{dt} = \omega p_a E_{\text{adult1}} - \delta_3 A_{\text{adult1}} + \gamma C_{\text{adult1}} - \mu_h A_{\text{adult1}}$$
$$\frac{dC_{\text{adult1}}}{dt} = \omega(1-p_a)(1-p_{\text{seek}})E_{\text{adult1}} - \gamma C_{\text{adult1}} - \mu_h C_{\text{adult1}}$$
$$\frac{dCt_{\text{adult1}}}{dt} = \omega(1-p_a)p_{\text{seek}}E_{\text{adult1}} - \tau Ct_{\text{adult1}} - \mu_h Ct_{\text{adult1}}$$
$$\frac{d\text{Sev}_{\text{adult1}}}{dt} = \nu C_{\text{adult1}} - (\epsilon + \tau_{\text{sev}} + \mu_{\text{sev1}} + \mu_h)\text{Sev}_{\text{adult1}}$$
$$\frac{dH_{\text{adult1}}}{dt} = \tau_{\text{sev}}\text{Sev}_{\text{adult1}} - (r_{\text{sev}} + \mu_{\text{sev2}} + \mu_h)H_{\text{adult1}}$$
$$\frac{dT_{\text{adult1}}}{dt} = \tau Ct_{\text{adult1}} - rT_{\text{adult1}} - \mu_h T_{\text{adult1}}$$
$$\frac{dR_{\text{adult1}}}{dt} = \delta_3 A_{\text{adult1}} + rT_{\text{adult1}} + r_{\text{sev}}H_{\text{adult1}} - \rho_3 R_{\text{adult1}} - \mu_h R_{\text{adult1}}$$

#### Adult2 with level 1 immunity Dynamics

$$\begin{aligned}
\frac{dE_{\text{adult2}}}{dt} &= \lambda_2 R_{\text{adult1}} - \omega E_{\text{adult2}} - r_{\text{iptp}} \cdot \text{prop}_{\text{iptp}} \cdot \text{prop}_{\text{pregnant}} \cdot E_{\text{adult2}} - \mu_h E_{\text{adult2}}, \\
\frac{dA_{\text{adult2}}}{dt} &= \omega P_{a2} E_{\text{adult2}} - \delta A_{\text{adult2}} + \gamma C_{\text{adult2}} - r_{\text{iptp}} \cdot \text{prop}_{\text{iptp}} \cdot \text{prop}_{\text{pregnant}} \cdot A_{\text{adult2}} - \mu_h A_{\text{adult2}}, \\
\frac{dC_{\text{adult2}}}{dt} &= \omega(1-P_{a2})(1-p_{\text{seek}})E_{\text{adult2}} - \gamma C_{\text{adult2}} - \mu_h C_{\text{adult2}} + \epsilon \cdot \text{Sev}_{\text{adult2}} - \nu C_{\text{adult2}}, \\
\frac{dCt_{\text{adult2}}}{dt} &= \omega(1-p_a)p_{\text{seek}}E_{\text{adult2}} - \tau C_{\text{t, adult2}} - \mu_h C_{\text{t, adult2}}, \\
\frac{d\text{Sev}_{\text{adult2}}}{dt} &= \nu C_{\text{adult2}} - (\epsilon + \tau_{\text{sev}} + \mu_{\text{sev1}} + \mu_h) \cdot \text{Sev}_{\text{adult2}}, \\
\frac{dH_{\text{adult2}}}{dt} &= \tau_{\text{sev}}\text{Sev}_{\text{adult2}} - (r_{\text{sev}} + \mu_{\text{sev2}} + \mu_h)H_{\text{adult2}}, \\
\frac{dT_{\text{adult2}}}{dt} &= \tau C_{\text{t, adult2}} - rT_{\text{adult2}} - \mu_h T_{\text{adult2}}, \\
\frac{dR_{\text{adult2}}}{dt} &= \delta A_{\text{adult2}} + rT_{\text{adult2}} - \lambda_2 R_{\text{adult2}} + \rho_3 R_{\text{adult3}} - \mu_h R_{\text{adult2}} - r_{\text{iptp}} \cdot \text{prop}_{\text{iptp}} \\
&\quad \cdot \text{prop}_{\text{pregnant}} \cdot R_{\text{adult2}} - \rho_2 R_{\text{adult2}} + \gamma_{\text{iptp}} P_{\text{iptp3}}, \\
\frac{dP_{\text{iptp2}}}{dt} &= r_{\text{iptp}} \cdot \text{prop}_{\text{iptp}} \cdot \text{prop}_{\text{pregnant}} \cdot (R_{\text{adult1}} + E_{\text{adult2}} + A_{\text{adult2}}) - \mu_h P_{\text{iptp2}} - \gamma_{\text{iptp}} P_{\text{iptp2}}.
\end{aligned}$$

#### Adult3 with level 2 immunity Dynamics

$$\begin{aligned}
\frac{dE_{\text{adult3}}}{dt} &= \lambda_2 R_{\text{adult2}} + \lambda_2 R_{\text{adult3}} - \omega E_{\text{adult3}} - r_{\text{iptp}} \cdot \text{prop}_{\text{iptp}} \cdot \text{prop}_{\text{pregnant}} \cdot E_{\text{adult3}} - \mu_h E_{\text{adult3}}, \\
\frac{dA_{\text{adult3}}}{dt} &= \omega E_{\text{adult3}} - \delta A_{\text{adult3}} - r_{\text{iptp}} \cdot \text{prop}_{\text{iptp}} \cdot \text{prop}_{\text{pregnant}} \cdot A_{\text{adult3}} - \mu_h A_{\text{adult3}}, \\
\frac{dR_{\text{adult3}}}{dt} &= \delta A_{\text{adult3}} - \rho_3 R_{\text{adult3}} - \mu_h R_{\text{adult3}} - r_{\text{iptp}} \cdot \text{prop}_{\text{iptp}} \cdot \text{prop}_{\text{pregnant}} \cdot R_{\text{adult3}} - \lambda_2 R_{\text{adult3}}, \\
\frac{dP_{\text{iptp3}}}{dt} &= r_{\text{iptp}} \cdot \text{prop}_{\text{iptp}} \cdot \text{prop}_{\text{pregnant}} \cdot (R_{\text{adult2}} + E_{\text{adult3}} + A_{\text{adult3}} + R_{\text{adult3}}) - \mu_h P_{\text{iptp3}} - \gamma_{\text{iptp}} P_{\text{iptp3}}.
\end{aligned}$$

### Appendix E Parameters used in the malaria model for Atacora, Alibori, Donga, and Borgou.

**Table E2**: Parameters used in the malaria model for Atacora, Alibori, Donga, and Borgou.

| Parameter | Meaning | Atacora | Alibori | Donga | Borgou | Source |
|---|---|---|---|---|---|---|
| mu_h | Human death rate | 2.28e-05 | 2.28e-05 | 2.28e-05 | 2.28e-05 | [14] |
| P | Population | 1115995.42 | 1058462.7 | 711058.43 | 1470961.98 | [14] |
| mu_b | Birth rate | 9.47e-05 | 9.47e-05 | 9.47e-05 | 9.47e-05 | [14] |
| rho_1 | Loss of immunity rate for level 1 immunity | 1/180 | 1/180 | 1/180 | 1/180 | Expert opinion (Supervisor) |
| rho_2 | Loss of immunity rate for level 2 immunity | 1/365 | 1/365 | 1/365 | 1/365 | Expert opinion (Supervisor) |
| rho_3 | Loss of immunity rate for level 3 immunity | 1/730 | 1/730 | 1/730 | 1/730 | Expert opinion (Supervisor) |
| Pa0 | Probability of asymptomatic (level 0) | 0.05 | 0.05 | 0.05 | 0.05 | Data fitting process |
| Pa1 | Probability of asymptomatic (level 1) | 0.3 | 0.3 | 0.3 | 0.3 | Data fitting process |
| Pa2 | Probability of asymptomatic (level 2) | 0.7 | 0.7 | 0.7 | 0.7 | Data fitting process |
| omega | Onset of infectiousness rate | 0.05 | 0.05 | 0.05 | 0.05 | [15] |
| p_a | Probability of asymptomatic for children | 0.2 | 0.2 | 0.2 | 0.2 | Data fitting process |
| r_smc | Rate of seasonal malaria chemoprevention | 1/30 | 1/30 | 1/30 | 1/30 | [16] |
| prop_smc | Proportion receiving SMC | 0.7 | 0.7 | 0 | 0 | NMCP of Benin |
| r_iptp | Rate of IPTp treatment | 1/30 | 1/30 | 1/30 | 1/30 | [2] |
| prop_iptp | Proportion of pregnant women on IPTp | 0.5 | 0.5 | 0.5 | 0.5 | [14] |
| peak | Seasonal forcing peak | 3 | 3 | 3 | 3 | [4] |
| itn_eff | ITN effectiveness | 0.5 | 0.5 | 0.5 | 0.5 | Data fitting process |
| irs_eff | IRS effectiveness | 0.65 | 0.65 | 0.65 | 0.65 | Data fitting process |
| gamma_iptp | Prophylaxis rate for IPTp | 1/210 | 1/210 | 1/210 | 1/210 | [2] |
| gamma_smc | Prophylaxis rate for SMC | 1/120 | 1/120 | 1/120 | 1/120 | [2] |
| alpha | Rate of aging to adults | 0.2/365 | 0.2/365 | 0.2/365 | 0.2/365 | [14] |
| prop_pregnant | Proportion of pregnant women | 0.000188 | 0.000188 | 0.000188 | 0.000188 | [14] |
| r | Recovery rate after treatment | 1/7 | 1/7 | 1/7 | 1/7 | [17] |
| zeta_a | Relative infectiousness (asymptomatic) | 0.25 | 0.25 | 0.25 | 0.25 | Data fitting process |
| delta | Recovery rate from asymptomatic | 0.01 | 0.01 | 0.01 | 0.01 | [17] |
| nu | Severe malaria rate | 1/20 | 1/20 | 1/20 | 1/20 | Data fitting process |

| Parameter | Meaning | Atacora | Alibori | Donga | Borgou | Source |
|---|---|---|---|---|---|---|
| epsilon | Recovery rate (severe malaria) | 1/20 | 1/20 | 1/20 | 1/20 | [18] |
| tau_sev | Treatment seeking rate (severe malaria) | 1/10 | 1/10 | 1/10 | 1/10 | [18] |
| r_sev | Recovery rate (severe malaria) | 1/5 | 1/5 | 1/5 | 1/5 | [18] |
| mu_sev1 | Death rate (untreated severe malaria) | 0.1/30 | 0.1/30 | 0.1/30 | 0.1/30 | Data fitting process |
| mu_sev2 | Death rate (treated severe malaria) | 0.15/50 | 0.15/50 | 0.15/50 | 0.15/50 | Data fitting process |
| a | Biting rate | 0.5 | 0.4 | 0.45 | 0.38 | Data fitting process |
| amp1 | Seasonal forcing amplitude (group 1) | 0.1 | 0.1 | 0.1 | 0.1 | [4] |
| phi1 | Seasonal forcing phase (group 1) | 2.5 | 2.5 | 2.5 | 2.5 | [4] |
| amp2 | Seasonal forcing amplitude (group 2) | 0.1 | 0.1 | 0.1 | 0.1 | [4] |
| phi2 | Seasonal forcing phase (group 2) | 6.5 | 6.5 | 6.5 | 6.5 | [4] |

### Appendix F Model Compartments and Their Definitions

**Table F3**: Model Compartments and Their Definitions

| Compartment | Definition |
|---|---|
| $dS\_under2$ | Susceptible under 2 |
| $dE\_under2$ | Exposed (latent) under 2 |
| $dA\_under2$ | Asymptomatic infectious under 2 |
| $dC\_under2$ | Clinical cases (not going to get treatment) under 2 |
| $dCt\_under2$ | Clinical cases (going to get treatment) under 2 |
| $dSev\_under2$ | Severe malaria cases under 2 |
| $dH\_under2$ | Hospitalized under 2 |
| $dT\_under2$ | Individuals receiving treatment under 2 |
| $dR\_under2$ | Recovered under 2 |
| $dP\_smc2$ | Individuals under SMC protection under 2 |
| $dS\_25$ | Susceptible aged 2–5 |
| $dE\_25$ | Exposed (latent) aged 2–5 |
| $dA\_25$ | Asymptomatic infectious aged 2–5 |
| $dC\_25$ | Clinical cases (not going to get treatment) aged 2–5 |
| $dCt\_25$ | Clinical cases (going to get treatment) aged 2–5 |
| $dSev\_25$ | Severe malaria cases aged 2–5 |
| $dH\_25$ | Hospitalized aged 2–5 |
| $dT\_25$ | Individuals receiving treatment aged 2–5 |
| $dR\_25$ | Recovered aged 2–5 |
| $dP\_smc25$ | Individuals under SMC protection aged 2–5 |
| $dS\_510$ | Susceptible aged 5–10 |
| $dE\_510$ | Exposed (latent) aged 5–10 |
| $dA\_510$ | Asymptomatic infectious aged 5–10 |
| $dC\_510$ | Clinical cases (not going to get treatment) aged 5–10 |
| $dCt\_510$ | Clinical cases (going to get treatment) aged 5–10 |
| $dSev\_510$ | Severe malaria cases aged 5–10 |
| $dH\_510$ | Hospitalized aged 5–10 |
| $dT\_510$ | Individuals receiving treatment aged 5–10 |
| $dR\_510$ | Recovered aged 5–10 |
| $dS\_adult$ | Susceptible adults (No immune protection assumed) |
| $dE\_adult$ | Exposed adults (No immune protection assumed) |

| Compartment | Definition |
|---|---|
| $dA\_adult$ | Asymptomatic infectious adults (No immune protection assumed) |
| $dC\_adult$ | Clinical cases (not going to get treatment) adults (No immune protection assumed) |
| $dCt\_adult$ | Clinical cases (going to get treatment) adults (No immune protection assumed) |
| $dSev\_adult$ | Severe malaria cases adults (No immune protection assumed) |
| $dH\_adult$ | Hospitalized adults (No immune protection assumed) |
| $dT\_adult$ | Individuals receiving treatment adults (No immune protection assumed) |
| $dR\_adult$ | Recovered adults (No immune protection assumed) |
| $dP\_iptp0$ | IPTp-protected adult females (No immune protection assumed) |
| $dE\_adult1$ | Exposed pregnant women (Level 1 immunity) |
| $dA\_adult1$ | Asymptomatic infectious (Level 1 immunity) |
| $dC\_adult1$ | Clinical cases (not going to get treatment) (Level 1 immunity) |
| $dCt\_adult1$ | Clinical cases (going to get treatment) (Level 1 immunity) |
| $dSev\_adult1$ | Severe cases (Level 1 immunity) |
| $dH\_adult1$ | Hospitalized (Level 1 immunity) |
| $dT\_adult1$ | Under treatment (Level 1 immunity) |
| $dR\_adult1$ | Recovered (Level 1 immunity) |
| $dP\_iptp1$ | IPTp-protected (Level 1 immunity) |
| $dE\_adult2$ | Exposed pregnant women (Level 2 immunity) |
| $dA\_adult2$ | Asymptomatic infectious (Level 2 immunity) |
| $dC\_adult2$ | Clinical cases (not going to get treatment) (Level 2 immunity) |
| $dCt\_adult2$ | Clinical cases (going to get treatment) (Level 2 immunity) |
| $dSev\_adult2$ | Severe cases (Level 2 immunity) |
| $dH\_adult2$ | Hospitalized (Level 2 immunity) |
| $dT\_adult2$ | Under treatment (Level 2 immunity) |
| $dR\_adult2$ | Recovered (Level 2 immunity) |
| $dP\_iptp2$ | IPTp-protected (Level 2 immunity) |
| $dE\_adult3$ | Exposed pregnant women (Level 3 immunity) |
| $dA\_adult3$ | Asymptomatic infectious (Level 3 immunity) |
| $dR\_adult3$ | Recovered (Level 3 immunity) |
| $dP\_iptp3$ | IPTp-protected (Level 3 immunity) |
